# Genome wide association study of clinical duration and age at onset of sporadic CJD

**DOI:** 10.1101/2023.10.17.23297050

**Authors:** Holger Hummerich, Helen Speedy, Tracy Campbell, Lee Darwent, Elizabeth Hill, Steven Collins, Christiane Stehmann, Gabor G Kovacs, Michael D Geschwind, Karl Frontzek, Herbert Budka, Ellen Gelpi, Adriano Aguzzi, Sven J van der Lee, Cornelia M van Duijn, Pawel P Liberski, Miguel Calero, Pascual Sanchez-Juan, Elodie Bouaziz-Amar, Jean-Louis Laplanche, Stéphane Haïk, Jean-Phillipe Brandel, Angela Mammana, Sabina Capellari, Anna Poleggi, Anna Ladogana, Maurizio Pocchiari, Saima Zafar, Stephanie Booth, Gerard H Jansen, Aušrinė Areškevičiūtė, Eva Løbner Lund, Katie Glisic, Piero Parchi, Peter Hermann, Inga Zerr, Brian S Appleby, John Collinge, Simon Mead

**Affiliations:** Medical Research Council Prion Unit, University College London Institute of Prion Diseases, London, UK; Australian National Creutzfeldt-Jakob Disease Registry, The Florey, Department of Medicine (RMH), The University of Melbourne, Victoria, 3010, Australia; Division of Neuropathology and Neurochemistry, Department of Neurology, Medical University of Vienna and Austrian Reference Center for Human Prion Diseases (ÖRPE), Vienna, Austria; Department of Laboratory Medicine and Pathobiology and Tanz Centre for Research in Neurodegenerative Disease, University of Toronto, and Laboratory Medicine Program & Krembil Brain Institute, University Health Network, Toronto, Ontario, Canada; UCSF Memory and Aging Center, Department of Neurology, University of California, San Francisco, USA; Institute of Neuropathology, University of Zürich, Zürich, Switzerland; Genomics of Neurodegenerative Diseases and Aging, Human Genetics, Vrije Universiteit Amsterdam, Amsterdam UMC, location VUmc, Amsterdam, The Netherlands; Alzheimer Center Amsterdam, Neurology, Vrije Universiteit Amsterdam, Amsterdam UMC location VUmc, Amsterdam, The Netherlands; Amsterdam Neuroscience, Neurodegeneration, Amsterdam, The Netherlands; Department of Epidemiology, Erasmus Medical Centre, Rotterdam, The Netherlands; Nuffield Department of Population Health, University of Oxford, UK; Department of Molecular Pathology and Neuropathology, Medical University of Lodz, Lodz, Poland; Chronic Disease Programme (UFIEC-CROSADIS) and Network Center for Biomedical Research in Neurodegenerative Diseases (CIBERNED), Instituto de Salud Carlos III, Madrid, Spain; Neurology Service, University Hospital Marqués de Valdecilla (University of Cantabria, CIBERNED and IDIVAL), Santander, Spain; Department of Biochemistry and Molecular Biology, Lariboisière Hospital, GHU AP-HP.Nord, University of Paris Cité, France; Sorbonne Université, INSERM, CNRS UMR 7225, Institut du Cerveau et de la Moelle épinière, ICM, Paris, France; Cellule nationale de référence des maladies de Creutzfeldt-Jakob, AP-HP, University Hospital Pitié-Salpêtrière, Paris, France; IRCCS, Istituto delle Scienze Neurologiche di Bologna, Bologna, Italy; Department of Biomedical and Neuromotor Sciences, University of Bologna, Bologna, Italy; Department of Cell Biology and Neuroscience, Istituto Superiore di Sanità, Rome, Italy; Department of Neurology, Clinical Dementia Center and National Reference Center for CJD Surveillance, University Medical School, Göttingen, Germany; Biomedical Engineering and Sciences Department, School of Mechanical and Manufacturing Engineering, National University of Sciences and Technology, Islamabad, Pakistan; Prion Disease Program, National Microbiology Laboratory, Public Health Agency of Canada, Winnipeg, Canada; Department of Pathology and Laboratory Medicine, University of Ottawa, Ottawa, Canada; Danish Reference Center for Prion Diseases, Department of Pathology, Copenhagen University Hospital, Rigshospitalet, Copenhagen 2100, Denmark; Department of Clinical Medicine, University of Copenhagen, Copenhagen, Denmark; National Prion Disease Pathology Surveillance Center, Case Western Reserve University, Cleveland, OH, USA; German Center for Neurodegenerative Diseases (DZNE), Göttingen, Germany

## Abstract

Human prion diseases are rare, transmissible and often rapidly progressive dementias. The most common type, sporadic Creutzfeldt-Jakob disease (sCJD), is highly variable in clinical duration and age at onset. Genetic determinants of late onset or slower progression might suggest new targets for research and therapeutics. We assembled and array genotyped sCJD cases diagnosed in life or at autopsy. Clinical duration (median:4, interquartile range (IQR):2.5-9 (months)) was available in 3,773 and age at onset (median:67, IQR:61-73 (years)) in 3,767 cases. Phenotypes were successfully transformed to approximate normal distributions allowing genome-wide analysis without statistical inflation. 53 SNPs achieved genome-wide significance for the clinical duration; all of which were located at chromosome 20 (top SNP rs1799990, pvalue=3.45×10^-36^, beta=0.34 for an additive model; rs1799990, pvalue=9.92×10^-67^, beta=0.84 for a heterozygous model). Fine mapping, conditional and expression analysis suggests that the well-known non-synonymous variant at codon 129 is the obvious outstanding genome-wide determinant of clinical duration. Pathway analysis and suggestive loci are described. No genome-wide significant SNP determinants of age at onset were found, but the *HS6ST3* gene was significant (pvalue=1.93 × 10^-6^) in a gene-based test. We found no evidence of genome-wide genetic correlation between case-control (disease risk factors) and case-only (determinants of phenotypes) studies. Relative to other common genetic variants, *PRNP* codon 129 is by far the outstanding modifier of CJD survival suggesting only modest or rare variant effects at other genetic loci.

## Introduction

Human prion diseases are rare and often rapidly progressive dementia disorders with no known treatments that slow the disease process. The most common type, sporadic Creutzfeldt-Jakob disease (sCJD), occurs at a relatively uniform annual incidence of 1-2/million/year, equating to a lifetime risk of approximately 1:5000(1). The clinical presentation and progression of the disorder is remarkably variable both in terms of the initial symptoms and signs, age at onset and clinical duration(2–4). Patients typically present in late middle or old age but have been reported in adolescence and early adulthood, and at the extremes of old age(5–7). The median clinical duration is usually reported as five months with a range of only a few weeks to several years(2). Ability to estimate the likely clinical duration could help with timely decisions about care(8).

Prions are proteinaceous pathogens formed of host prion protein (PrP) which cause mammalian prion diseases like bovine spongiform encephalopathy, sheep scrapie, chronic wasting disease of cervids, and the human disorders(9). The recently determined structures of mouse and hamster prions reveals assemblies of PrP in a parallel in-register beta sheet structure with two domains(10, 11), in marked contrast to the predominant alpha-helices of normal cellular PrP(12). Prions are thought to replicate by a process of binding of normal cellular PrP, conformational change and subsequently aggregate fission. In several model systems, incubation time of prion disease is influenced by PrP gene expression, primary sequence and polymorphisms, as well as prion strains(13), thought to be conferred by structural variation of the pathogen(14). Experiments using animal or cellular model systems have led to proposals of several possible non-PrP mechanisms of toxicity in prion diseases, involving PrP binding partners on the cell surface and downstream intracellular changes(15–17); however, their relevance to the human diseases is yet to be determined.

Human epidemiological and genetic studies have identified factors that associate with survival time in sCJD(2, 8, 18, 19), including demography, prion protein genotype, molecular strain typing of protease-resistant prion protein by Western blot analysis, and a range of biofluid, tissue, imaging, and neurophysiological biomarkers(20). Many biomarkers simply measure the rate or extent of neuronal injury, loss, or dysfunction, or immune cell or glial responses, whereas genetic associations are implicitly causal of modified clinical phenotypes. In this study, we sought to determine the effects of genome-wide common genetic variation on key clinical phenotypes of sCJD, to develop evidence of modifiers relevant to human prion diseases that might benefit understanding of disease processes and generate new ideas for therapeutics.

## Methods

### Diagnosis and clinical phenotypes

Details of the contributing sites and diagnostic criteria were given in a previous publication(19). In short, all patient participants were deceased and gained a diagnosis in life of probable CJD or definite CJD after a post-mortem examination (using contemporary epidemiological criteria which changed over the recruitment period 1990–2019). “Probable CJD” is an epidemiological term that equates to an almost certain diagnosis of CJD post-mortem (eg.(21)). Age at clinical onset was given to the nearest month. Clinical duration was based on the examining physician’s impression of the date of onset of the first symptom that subsequently was thought to be a component of the disease syndrome until death in months. Research ethics approval was obtained from the London-Harrow Research Ethics Committee.

### Genotyping and quality control

In addition to 4110 samples previously reported, genotyped on an Illumina OmniExpress array(19), 819 new samples were genotyped using Illumina’s Global Screening Array. Standard sample and genotyping quality control was performed using PLINK v1.90b3v, which generated 6,308,901 autosomal SNPs of high quality. Samples with a call rate below 98% and population outliers identified via multidimensional scaling were removed.

Additionally, related samples (Pi_Hat > 0.1875) were discarded. Only autosomal SNPs with a genotyping rate of >99%, a minor allele frequency ≥ 0.01 and SNPs not deviating from the Hardy-Weinberg equilibrium (P>10^-4^) were retained. SNPs of A/T or G/C transversion or those which showed deviation from heterozygosity mean (±3 SD) were excluded. To ensure consistency with the Michigan Imputation Server pipeline the target VCF files were checked against the 1000 Genomes Project reference panel (https://faculty.washington.edu/browning/conform-gt.html/). Genotypes were imputed using the Michigan Imputation Server (using Minimac4 assuming a mixed population, HRC r1.1 2016 as reference panel and Eagle 2.4 for phasing)(22). A post-imputation QC analysis was carried out and SNPs with an r^2^ threshold lower than 0.3 (removing 70% of poorly imputed SNPs) were excluded.

### Statistical analysis

SNPTEST (v2.5.2) was used to perform association and conditional analysis with an additive and heterozygous logistic regression model, using sex, contributing site and 10 population covariates generated with PLINK (v1.90b3v; www.cog-genomics.org/plink/1.9/). Genetic correlation between this (using duration as phenotype) and the previously conducted sCJD case-control study(19) was performed using LDSC(23), a software tool for LD score and heritability estimation using summary statistics. Meta-analysis was performed using METAL(22) combining the previously published GWAS case-control data(19) and the case-only data described here using summary test statistics as input (6,314,883 SNPs in the union list) and adopting the sample-based approach by combining z-scores across samples in a weighted sum proportional to study sample sizes. FUMA(24), using an integrated Magma gene-based and gene-set analysis on the GWAS summary data, was utilised to perform pathway analysis to identify genes and pathways associated with sCJD risk. FUMA also provides information about chromatin interaction, expression patterns and shared molecular functions between genes. MAGMA software was also utilised for gene-based / gene-set analysis(25). Power analysis was performed using R functions taken from the Github site kaustubhad/gwas-power(25).

## Results

We assembled 3999 (age: 3993; duration: 3971) cases of probable or definite sCJD by contemporary diagnostic criteria either included in a previous paper from the collaborative group(19), or newly genotyped on Illumina’s Global Screening Array (Table 1). Genotype doses were imputed using the Michigan Imputation Server(22), resulting in 6,308,901 SNPs passing quality control. All patients were deceased. Clinical duration (median:4.0, IQR:2.5-9 (months)) and age at onset (median:67, IQR:61-73 (years)) were available in 3773 cases.

**Table 1.** Number of collected samples from 12 countries (age / duration) with interquartile range and median.

| Country | N (age) | N (duration) | N (PrP type) | Median (age) | IQR (age) | Median (duration) | IQR (duration) |
| --- | --- | --- | --- | --- | --- | --- | --- |
| Australia | 22 | 22 | 22 | 66.6 | 12.52 | 2.05 | 1.87 |
| Austria | 44 | 44 | 40 | 72 | 7 | 4.5 | 5.87 |
| Canada | 135 | 135 | 0 | 67 | 14.5 | 4 | 5 |
| France | 96 | 96 | 49 | 67.85 | 12.9 | 4 | 4 |
| Germany | 792 | 798 | 409 | 66 | 12.22 | 6 | 8 |
| Italy | 554 | 554 | 424 | 67 | 13 | 5 | 7 |
| Netherlands | 126 | 126 | 0 | 66.51 | 13.44 | 3.94 | 4.02 |
| Poland | 42 | 42 | 0 | 63.5 | 9.25 | 3 | 2.88 |
| Spain | 74 | 71 | 0 | 69 | 13.75 | 3.45 | 4.25 |
| Switzerland | 35 | 35 | 32 | 69.56 | 13.48 | 2.69 | 2.91 |
| UK | 970 | 966 | 0 | 67 | 12 | 5 | 7.06 |
| USA | 1103 | 1082 | 213 | 67 | 12.17 | 4 | 6 |
| Total | 3993 | 3971 | 1189 |  |  |  |  |

The median age / duration for men was 67 years and 3.8 months respectively and 67 years and 4.0 months for women. Median clinical duration (2.0-6.0 months) and age at onset (63.5-72 years) varied by site, so this was included as a covariate in the analysis. Phenotypes were modelled as normally distributed quantitative traits following transformation using methods developed by Box and Cox(26). Association analysis omitting sex, age or country or any combination as covariates did not show any significant difference in terms of outcome. Principal components analysis was used to exclude cases with distinct ancestry (n=54) and did not suggest any strong effects of ancestry on the outcomes of interest (Supplementary Figures 2, 3).

Additive and heterozygous genetic models were run genome-wide in SNPTEST with sex, contributing site and genetic ancestry covariates (see Methods) without any statistical inflation (lambda=1.000 / 1.000 for clinical duration / age) as illustrated with QQ plots in Figure 3a (duration phenotype) and 3b (age phenotype). 53 SNPs achieved genome-wide significance (P<5×10^-8^) for the clinical duration phenotype (additive model) (Figure 3c; Supplementary Table 1a), all at the *PRNP* locus (top SNP rs1799990, pvalue=3.45×10^-36^, beta=0.34 for additive model; rs1799990, pvalue=9.92×10^-67^, beta=0.84 for heterozygous model, Figures 4a,4b). *PRNP* rs1799990 was the obvious outstanding genome-wide candidate determinant of clinical duration. Of 68 cis-eQTL SNPs associated with *PRNP* expression in various brain tissues (obtained from GTEx); none were present in the list of 53 SNPs achieving genome-wide significance (duration phenotype). 50 of the eQTL SNPs for *PRNP* passed QC, all were P>0.001 (duration phenotype). No genome-wide significant SNPs remained after conditioning for rs1799990 codon 129 (Figure 4c). There were 51 suggestive associated SNPs (5×10^-8^ > pvalue<1×10^-5^, including at regions near to *HDHD5* (chromosome 22), *FHIT* (chromosome 3) and *EREG* (chromosome 4) (see Supplementary Table 1b; Supplementary Figures 5-7). There were no significant gene-based associations of clinical duration (MAGMA and FUMA, Table 2).

**Fig 1.**
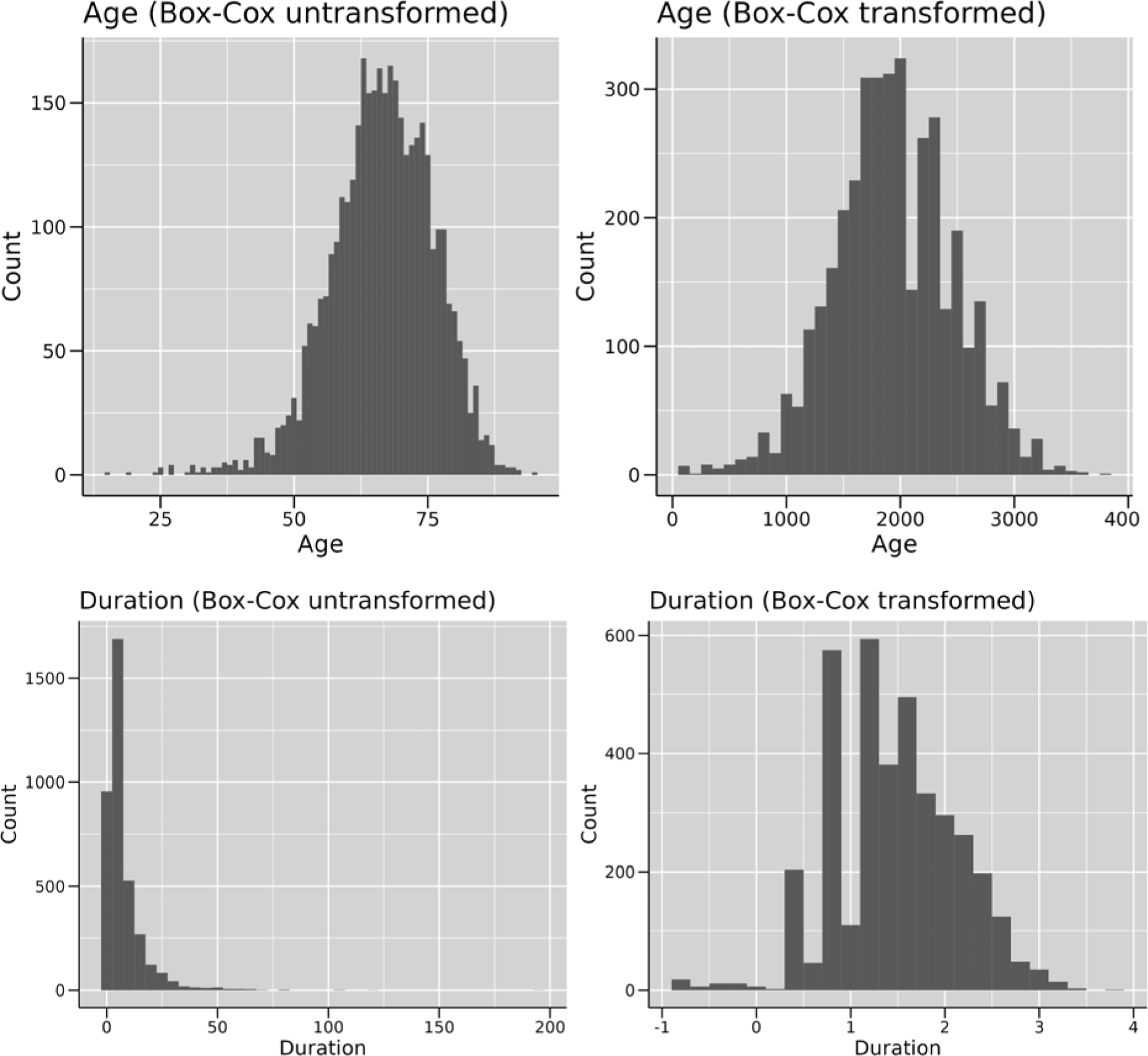
Histograms for phenotypes age before (A) and after (B) Box-Cox transformation and duration before (C) and after (D) Box-Cox transformation.

**Fig 2.**
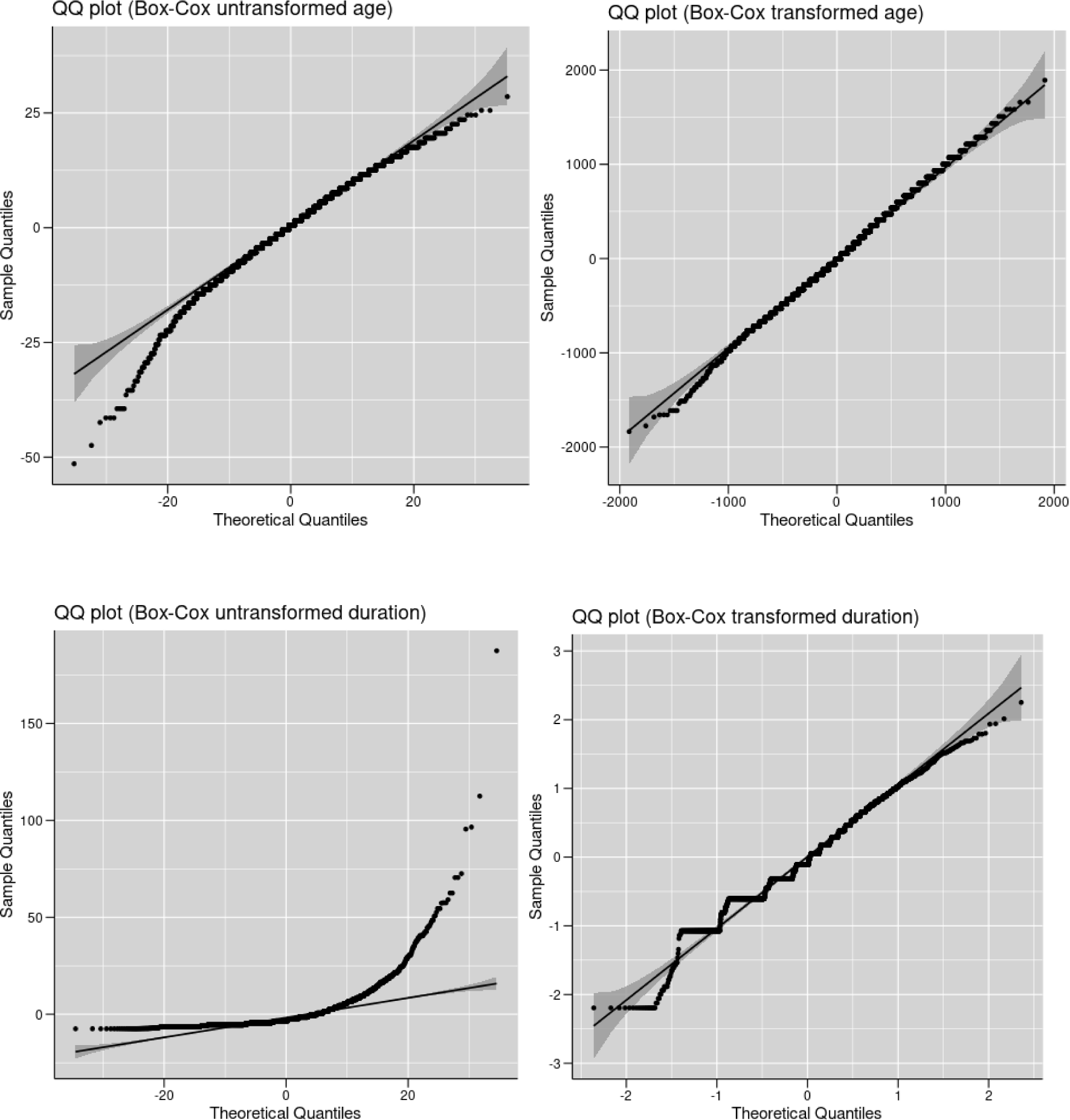
Quantile-Quantile plots for phenotypes age before (A) and after (B) Box-Cox transformation and duration before (C) and after (D) Box-Cox transformation.

**Figure 3a.**
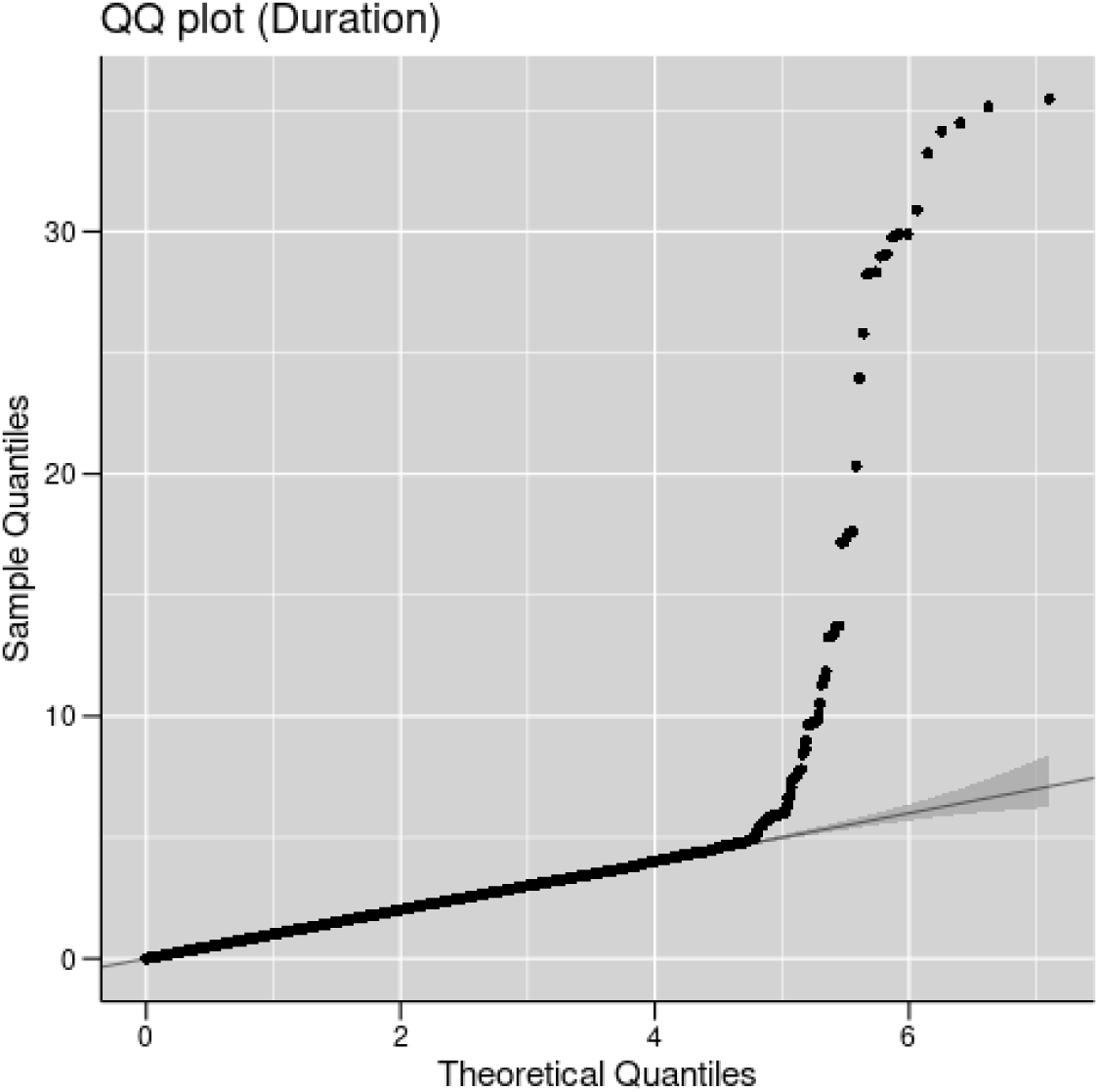
Quantile-Quantile plot with duration as phenotype.

**Figure 3b.**
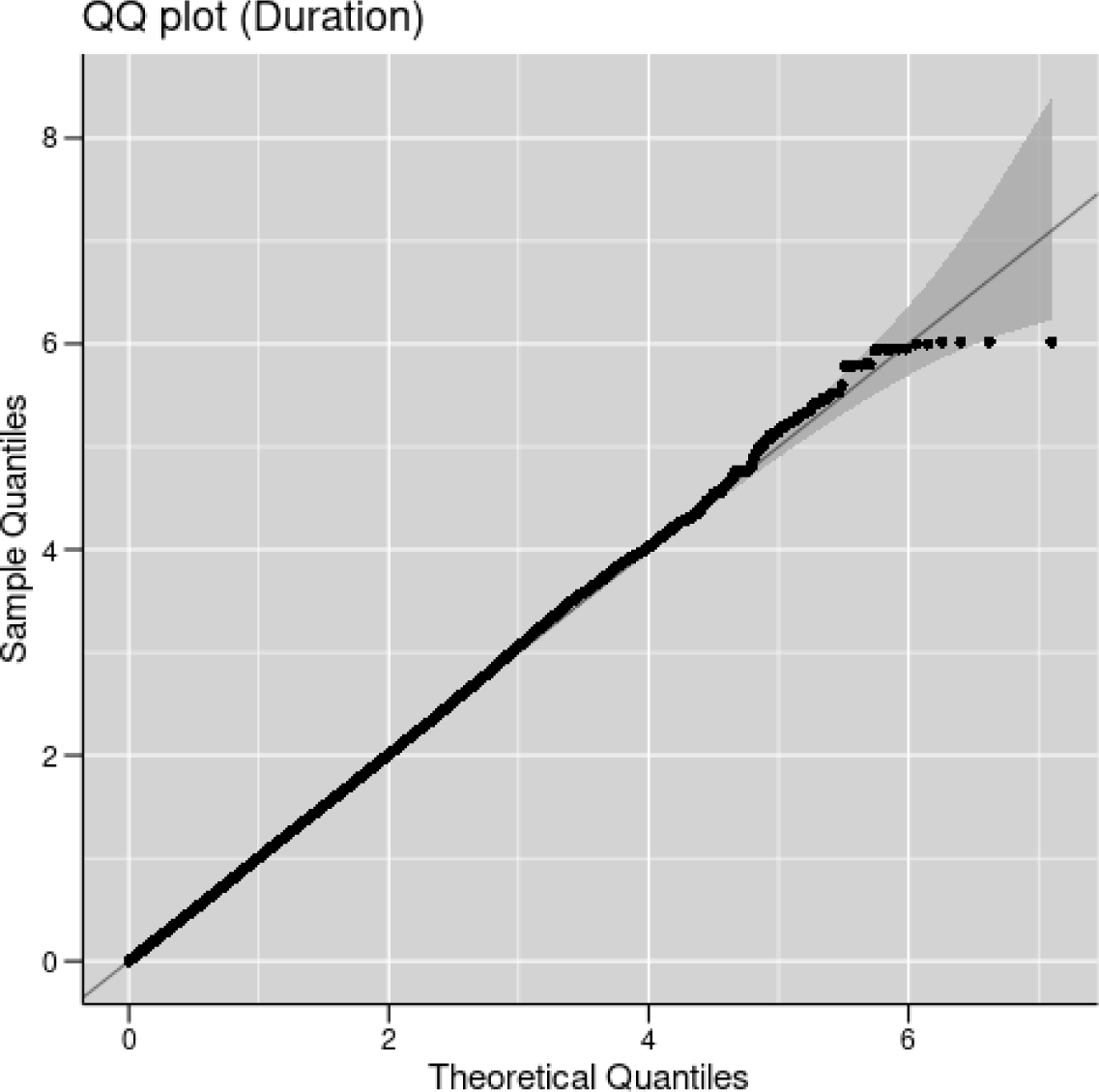
Quantile-Quantile plot with age as phenotype.

**Figure 3c.**
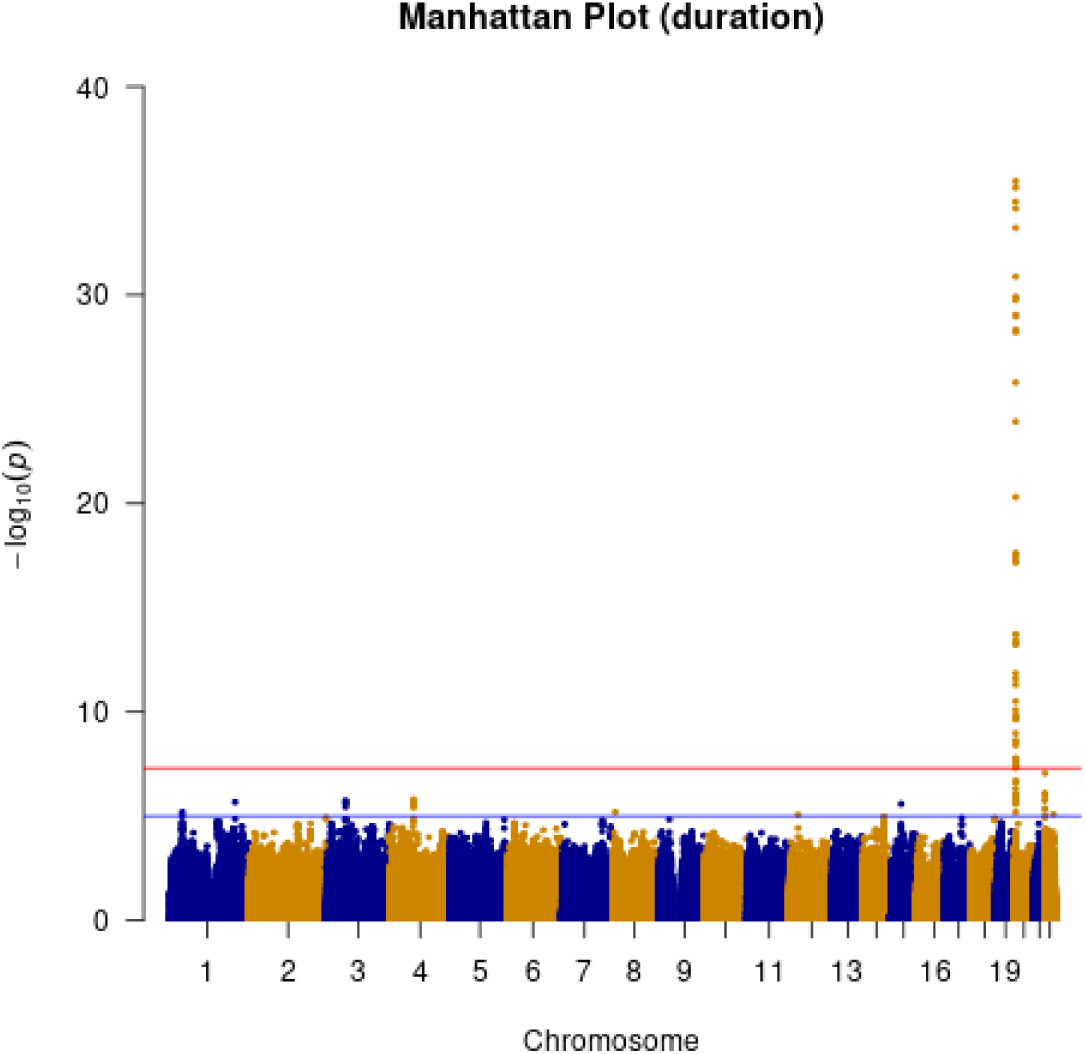
Manhattan plot with clinical duration as phenotype. (red line indicating genome-wide significance of 5×10^-8^; blue line indicating suggestive genome-wide significance (5×10^-8^ > pvalue < 1×10^-5^))

**Figure 3d.**
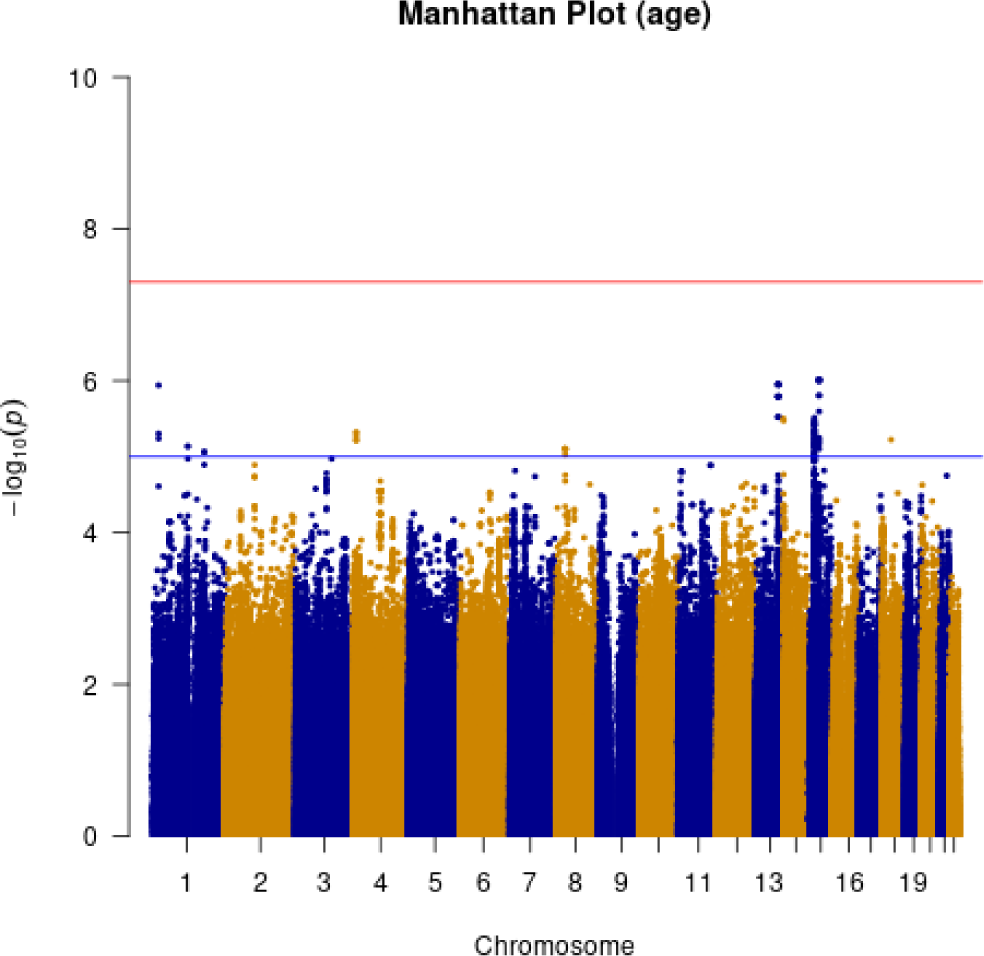
Manhattan plot with age at onset as phenotype. (red line indicating genome-wide significance of 5×10^-8^; blue line indicating suggestive genome-wide significance (5×10^-8^ > pvalue < 1×10^-^)

**Figure 4a.**
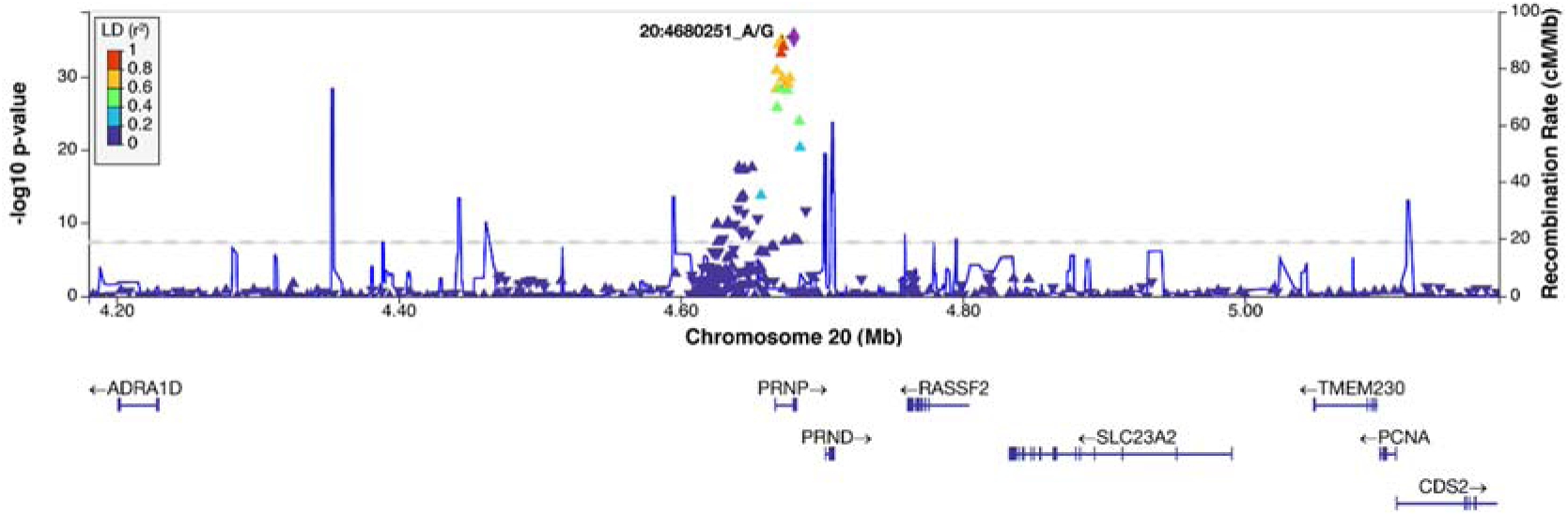
Regional association plot at *PRNP* locus with clinical duration as phenotype (additive model)

**Figure 4b.**
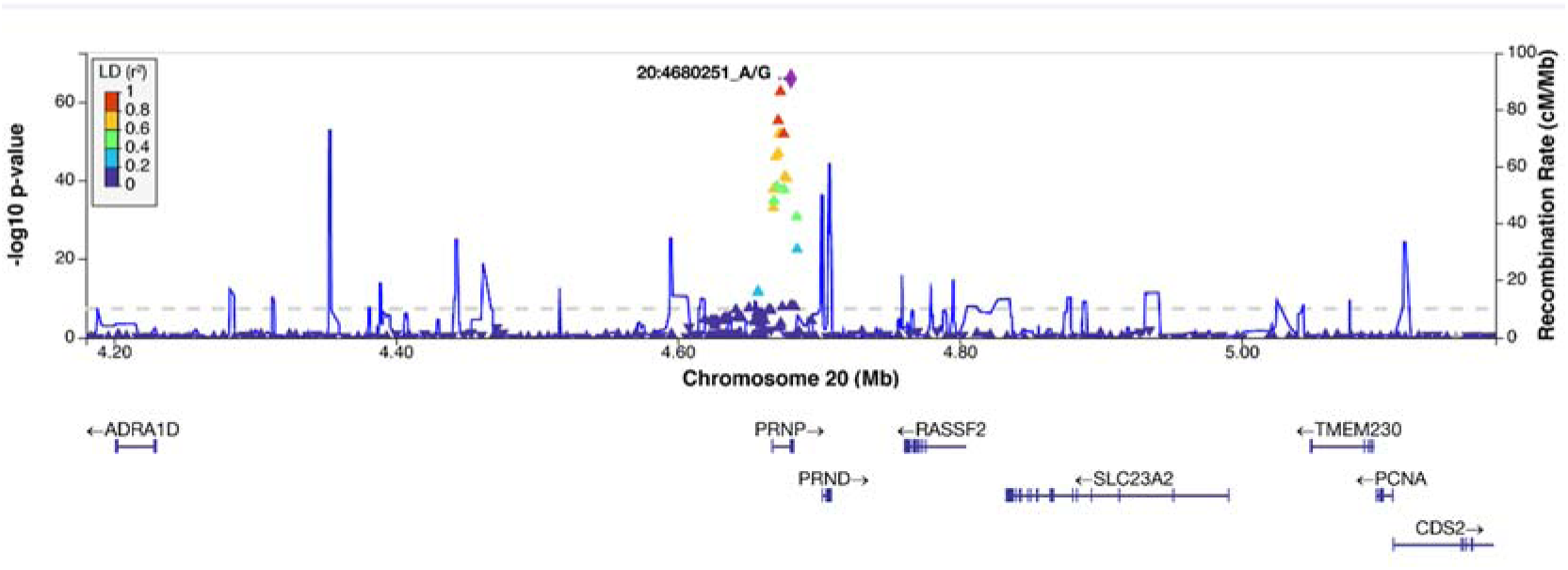
Regional association plot at *PRNP* locus with clinical duration as phenotype (heterozygous model)

**Figure 4c.**
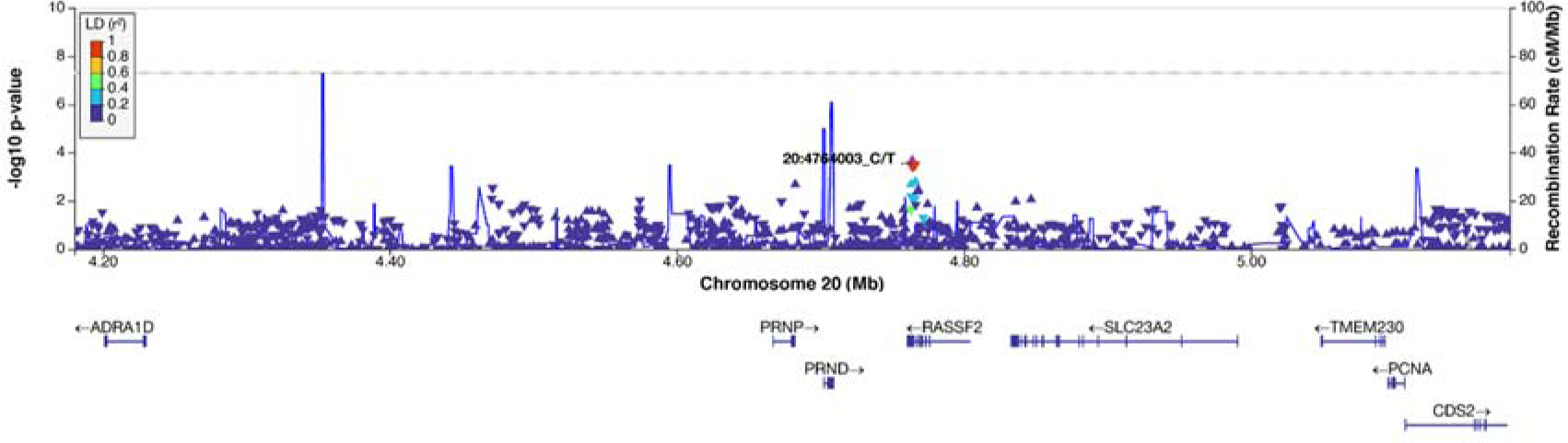
Regional association plot at *PRNP* locus for conditional analysis on SNP rs1799990 with clinical duration as phenotype (additive model)

**Table 2a.**
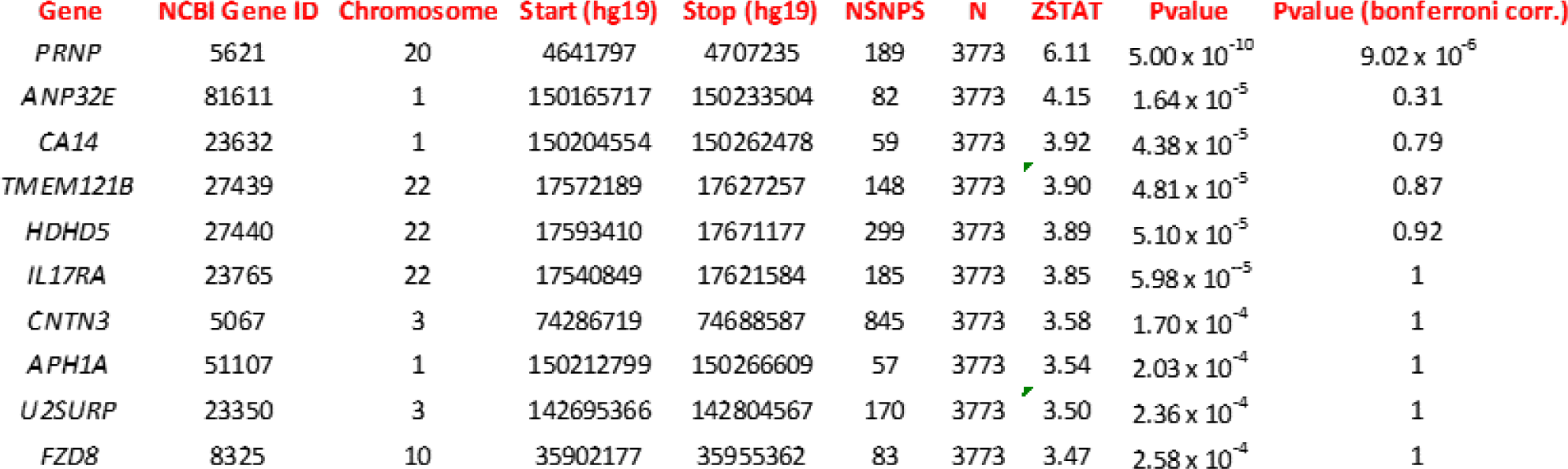
Top 10 genes identified by MAGMA (standalone) gene analysis (including genome-wide significant SNPs) with duration as phenotype. (NSNPS = number of SNPs annotated to a gene; N = number of samples; ZSTAT = Z-score for the gene, based on its p-value)

**Table 2b.** Top 10 genes identified by FUMA gene analysis (including genome-wide significant SNPs) with duration as phenotype. (NSNPS = number of SNPs annotated to a gene; N = number of samples; ZSTAT = Z-score for the gene, based on its p-value)

| GENE | CHR | START (hg19) | STOP (hg19) | NSNPS | N | ZSTAT | Pvalue | Pvalue (bonferroni corr.) |
| --- | --- | --- | --- | --- | --- | --- | --- | --- |
| PRNP | 20 | 4666882 | 4682236 | 27 | 3773 | 7.02 | $1.12 \times 10^{-12}$ | $2.03 \times 10^{-8}$ |
| CECR5 | 22 | 17618401 | 17646177 | 81 | 3773 | 4.28 | $9.42 \times 10^{-6}$ | 0.17 |
| ANP32E | 1 | 150190717 | 150208504 | 19 | 3773 | 3.86 | $5.63 \times 10^{-5}$ | 1 |
| CA14 | 1 | 150229554 | 150237478 | 3 | 3773 | 3.79 | $7.60 \times 10^{-5}$ | 1 |
| AL356356.1 | 1 | 150521897 | 150524367 | 1 | 3773 | 3.67 | $1.23 \times 10^{-4}$ | 1 |
| AC006946.15 | 22 | 17602476 | 17612994 | 40 | 3773 | 3.51 | $2.20 \times 10^{-4}$ | 1 |
| CCDC174 | 3 | 14693271 | 14714166 | 66 | 3773 | 3.51 | $2.26 \times 10^{-4}$ | 1 |
| KCNJ3 | 2 | 155554811 | 155714863 | 453 | 3773 | 3.45 | $2.82 \times 10^{-4}$ | 1 |
| VRK3 | 19 | 50479724 | 50529203 | 115 | 3773 | 3.38 | $3.56 \times 10^{-4}$ | 1 |
| U2SURP | 3 | 142683339 | 142779567 | 154 | 3773 | 3.38 | $3.61 \times 10^{-4}$ | 1 |

**Table 2c.**
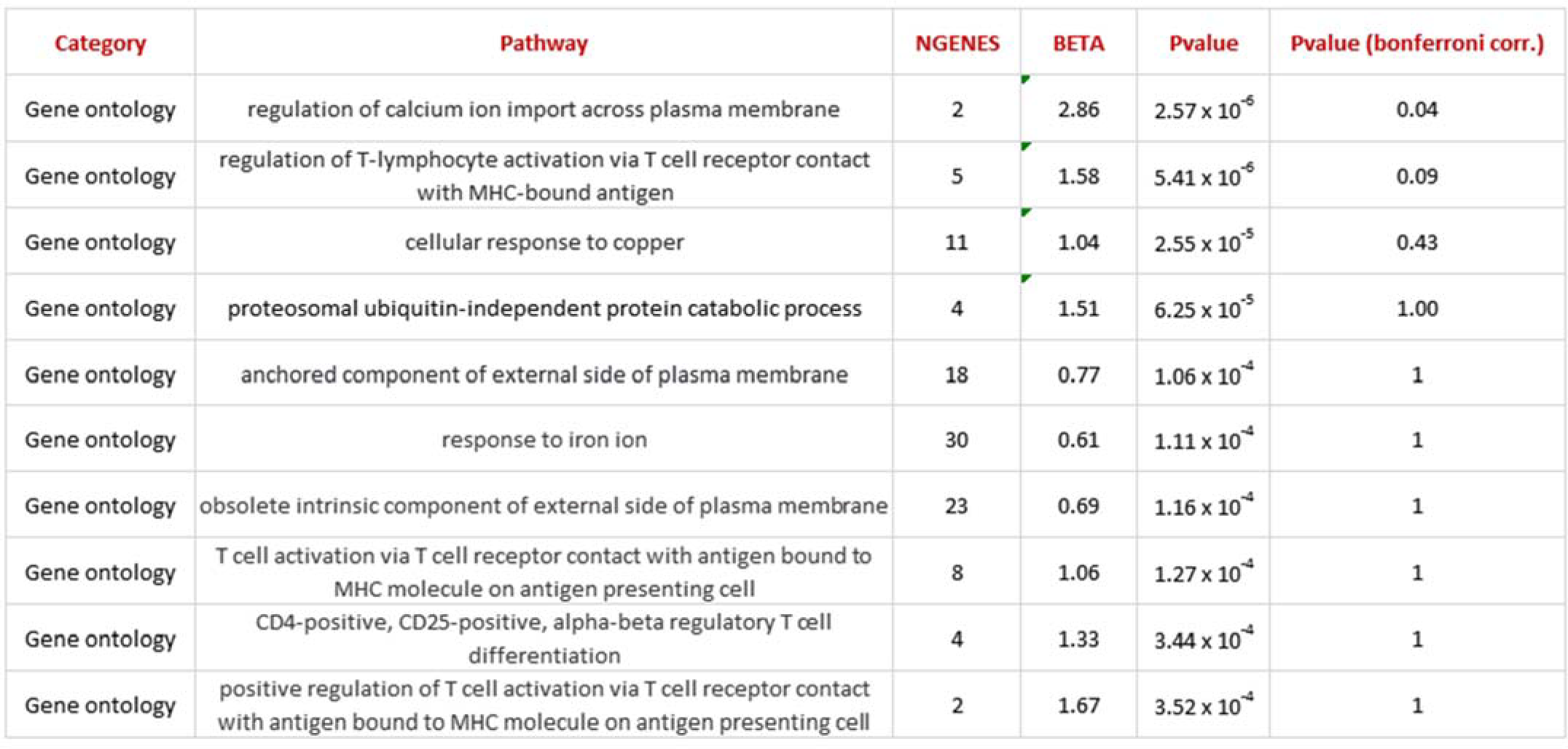
Top 10 pathways identified by MAGMA (standalone) gene-set analysis (including genome-wide SNPs) using duration as phenotype. (NGENES = number of genes in the gene-set dataset; BETA = regression coefficient of the gene set)

| Category | Pathway | NGENES | BETA | Pvalue | Pvalue (bonferroni corr.) |
| --- | --- | --- | --- | --- | --- |
| Gene ontology | regulation of calcium ion import across plasma membrane | 2 | 2.86 | $2.57 \times 10^{-6}$ | 0.04 |
| Gene ontology | regulation of T-lymphocyte activation via T cell receptor contact with MHC-bound antigen | 5 | 1.58 | $5.41 \times 10^{-6}$ | 0.09 |
| Gene ontology | cellular response to copper | 11 | 1.04 | $2.55 \times 10^{-5}$ | 0.43 |
| Gene ontology | proteosomal ubiquitin-independent protein catabolic process | 4 | 1.51 | $6.25 \times 10^{-5}$ | 1.00 |
| Gene ontology | anchored component of external side of plasma membrane | 18 | 0.77 | $1.06 \times 10^{-4}$ | 1 |
| Gene ontology | response to iron ion | 30 | 0.61 | $1.11 \times 10^{-4}$ | 1 |
| Gene ontology | obsolete intrinsic component of external side of plasma membrane | 23 | 0.69 | $1.16 \times 10^{-4}$ | 1 |
| Gene ontology | T cell activation via T cell receptor contact with antigen bound to MHC molecule on antigen presenting cell | 8 | 1.06 | $1.27 \times 10^{-4}$ | 1 |
| Gene ontology | CD4-positive, CD25-positive, alpha-beta regulatory T cell differentiation | 4 | 1.33 | $3.44 \times 10^{-4}$ | 1 |
| Gene ontology | positive regulation of T cell activation via T cell receptor contact with antigen bound to MHC molecule on antigen presenting cell | 2 | 1.67 | $3.52 \times 10^{-4}$ | 1 |

Age-based analysis did not identify any genome-wide significant SNP associations (Figure 3d). Two suggestive associations were identified on chromosome 15 near *NEDD4* and chromosome 13 near *UGGT2* (Supplementary Figures 9, 10). Gene-based analysis for age at onset with MAGMA identified *HS6ST3* (pvalue=1.93 × 10^-6^), with similarly significant association detected using FUMA (Supplementary Table 5).

Gene-set analysis for clinical duration (including *PRNP* locus) identified binders of type-5 metabotropic glutamate receptors (GO Molecular Function ontology n=1738, pvalue=1.97×10^-5^) (Table 2d). Gene-set analysis for age at onset revealed intracellular oxygen homeostasis as a significant term (pvalue=0.03, pvalue=1.89 × 10^-6^). Genetic correlation between clinical duration GWAS and the previously published case-control GWAS resulted in a non-significant genetic correlation of 0.1467 (pvalue=0.79, 95% CI 0.92,1.21; Supplementary Table 2). Meta-analysis of the two GWAS (case-only and case-control) resulted in the same strong codon 129 effect as described above whilst removing the suggestive locus on chromosome 22 the *HDHD5* locus (Supplementary Figure 8).

**Table 2d.** Top 10 pathways identified by FUMA gene-set analysis (including genome-wide SNPs) using duration as phenotype. (NGENES = number of genes in the gene-set dataset; BETA = regression coefficient of the gene set)

| Category | Pathway | NGENES | BETA | Pvalue | Pvalue (bonferroni corr) |
| --- | --- | --- | --- | --- | --- |
| Gene ontology | type_5_metabotropic_glutamate_receptor_binding | 5 | 1.92 | $1.85 \times 10^{-5}$ | 0.29 |
| Gene ontology | ureteric_bud_elongation | 9 | 1.00 | $8.58 \times 10^{-5}$ | 1 |
| Gene ontology | negative_regulation_of_cell_maturation | 8 | 0.89 | $1.53 \times 10^{-5}$ | 1 |
| Gene ontology | mechanosensory_behavior | 13 | 0.85 | $1.55 \times 10^{-5}$ | 1 |
| Gene ontology | actin_filament_based_transport | 8 | 0.90 | $3.84 \times 10^{-5}$ | 1 |
| Gene ontology | learned_vocalization_behavior_or_vocal_learning | 8 | 0.97 | $3.99 \times 10^{-5}$ | 1 |
| Gene ontology | peptidyltransferase_activity | 3 | 1.70 | $5.78 \times 10^{-5}$ | 1 |
| Gene ontology | pyrimidine_containing_compound_transmembrane_transport | 10 | 0.79 | $7.38 \times 10^{-5}$ | 1 |
| Curated gene sets | smid_breast_cancer_relapse_in_pleura_dn | 24 | 0.49 | $7.89 \times 10^{-5}$ | 1 |
| Gene ontology | vitamin_binding | 128 | 0.23 | $8.75 \times 10^{-4}$ | 1 |

We also calculated the power of the study based on 3773 samples and a genome-wide significance level of 5×10^-8^ using the additive model with a range of effect sizes and minor allele frequencies. Plotting the most significant SNP (*PRNP*; rs1799990) and the lead SNPs of the suggestive association signals (*HDHD5*, rs4819962; *FHIT*, rs2366847; *EREG*, rs11727991) resulted in rs1799990 achieving full power and the three lead SNPs being borderline achieving a power value of ∼0.7-0.8 (Supplementary Figure 11).

Interestingly, there was no evidence that the sCJD genetic susceptibility genes, *STX6* or *GAL3ST1,* which were identified in the previously published case-control study(19), modify clinical phenotypes. The identification of these genes in the case-control GWAS implicated intracellular trafficking and sphingolipid metabolism respectively as causal disease mechanisms. To further investigate the roles of these pathways in disease phenotypes, we compiled a comprehensive, bespoke gene list including genes related to these pathways, which have been implicated in neurodegenerative diseases, and performed MAGMA analysis (Supplementary Tables 3, 4). This highlighted *UGGT2,* a sphingolipid metabolism linked gene, to be associated with sCJD age of onset.

## Discussion

We describe the first well-powered GWAS for phenotypic traits in sporadic human prion disease. The only clearly identified risk locus was the *PRNP* gene itself, more specifically the well-known common variant at codon 129, for the clinical duration phenotype. Conditioning for the codon 129 polymorphism at this locus removed all evidence of association at the locus, implicating the coding sequence of *PRNP* and not PrP expression in controlling this phenotype. We found a number of suggestive risk loci with P<10^-5^, which will require additional genetic evidence before being considered further. Pathway analysis identified binders of type-5 metabotropic glutamate receptors, which are known to mediate the downstream effects of amyloid beta bound to prion protein, as a top hit for clinical duration (27, 28). Importantly however, since this small gene set (n=5) was non-significant after removing *PRNP*, these data should be interpreted with caution. Overall, this work further establishes the key importance of the PrP coding sequence relative to other potential mechanisms and genetic loci in determination of CJD survival.

For age at onset there were no genome-wide significant SNPs, but we identified the *HS6ST3* in a gene-based test and intracellular oxygen homeostasis by pathway analysis. *HS6ST3* or Heparan Sulfate 6-O-Sulfotransferase 3 catalyses the transfer of sulfate from 3’-phosphoadenosine 5’-phosphosulfate (PAPS) to position 6 of the N-sulfoglucosamine residue (GlcNS) of heparan sulfate (HS), thus potentially modifying the interactions of this molecule with cell surface proteins. There is a vast literature on a role for polyanionic compounds, including HS in prion disease pathogenesis, as they colocalise with PrP^C^ on the cell surface and with aggregated PrP^Sc^(29), act as potential co-factors in prion replication, and there is potent inhibitory activity of HS and related compounds on prion propagation(30). A role for intracellular oxygen homeostasis is less clearly linked to prion disease. Both associations were borderline in significance taking into account multiple testing.

We found no evidence of genetic correlation between the case-only and published case-control GWAS analyses. We observed only a moderate heritability (h^2^_SNP_=0.18-0ꞏ26, using different methods) for the case-control GWAS(19), and low heritability for the duration phenotype (h^2^_SNP_=0.09 using LDSC). Common SNPs measured in these studies therefore explain only a small proportion of disease phenotypes. The only locus common to both GWAS studies is *PRNP*, with no evidence that SNPs at the *STX6* or *GAL3ST1* loci have any effect on clinical phenotypes in lead SNP association, gene-based or pathway analyses. It is possible that larger sample sizes, with additional risk factor discovery, will uncover shared determinants, but the current evidence suggests that beyond *PRNP*, distinct mechanisms and/or stochasticity determines disease risk, age at onset and clinical duration.

Absence of an association between *PRNP* cis-eQTL SNPs and clinical duration/age of onset should not deter the pursuit of methods to reduce PrP as a therapeutic strategy. There is a wealth of evidence for the safety and potential effectiveness of this approach from animal models(31–35). *PRNP* cis-eQTL SNPs are predominantly associated with localised tissue expression of PrP, typically in cerebellum or cerebellar hemispheres, and are relatively modest effects. Therapeutic strategies aim for more profound protein knock-down, which will be critical to achieve across a wide range of central nervous system tissues and cell types(36).

It was imperative to transform the non-normal distribution of the duration phenotype data as the GWAS association model requires Gaussian distributed phenotype data to avoid model misspecification, which could lead to false conclusions. A number of data transformations were tested (log, rank inverse, square root) for transformation of the phenotype data (duration and age) and the Box-Cox transformation was found to be the best option for establishing the optimal correlation coefficient ensuring a normal distribution and reduction of data noise to a minimum.

This study was limited by sample size and was restricted to the examination of age at onset and clinical duration phenotypes that are almost universally collected, whereas the diversity of clinical phenotypes in CJD is well known (including variable involvement of cognitive, ataxic, psychiatric, sleep and motor aspects). In biochemical aspects and biomarkers, we see diversity of PrP^Sc^ types, and different imaging, neurophysiological and fluid biomarker associations. These parameters are only collected in smaller subsets of data. Genetic studies in a rare disease like sCJD benefit from national investment and collaboration in prion disease surveillance(37). Future work of the collaborative group might focus on building larger sample collections for increased power, exome or genome studies to ascertain rare and structural variants and extension of these type of analyses to other phenotypes (e.g., the well-known subtypes of CJD based on major symptom at presentation (ataxia, visual processing disorder etc.)).

## Data Availability

Summary statistics are available through the GWAS catalog at NHGRI-EBI via study accession number GCST90295967. Further data available upon request.

## Acknowledgements

We thank Richard Newton for support with images and UCL Genomics who did the array processing. For UK samples we would like to thank patients, their families and carers, UK neurologists and other referring physicians, co-workers at the National Prion Clinic, our colleagues at the National Creutzfeldt-Jakob Disease Research and Surveillance Unit, Edinburgh. Several authors at UCL/UCLH receive funding from the Department of Health’s NIHR Biomedical Research Centres funding scheme. Some of this work was supported by the Department of Health funded National Prion Monitoring Cohort study. Funding for the collection of Polish samples for study was partially provided by the EU joint programme JPND and Medical University of Lodz. We thank Dr. Maria Styczynska from Mossakowski Medical Research Centre; Polish Academy of Sciences; Warsaw, for kindly providing control DNA samples for the Polish cohort. The Italian national surveillance of Creutzfeldt-Jakob disease and related disorders is partially supported by the Ministero della Salute, Italy. The German National Reference Centre for TSE is funded by grants from the Robert-Koch-Institute. The Dutch National Prion Disease Registry is funded by the National Institute for Public Health and the Environment (RIVM), which is part from the Ministry for Health, Welfare and Sports, The Netherlands. PS-J was supported by Instituto de Salud Carlos III [Fondo de Investigación Sanitaria, PI16/01652] Accion Estrategica en Salud integrated in the Spanish National I+D+i Plan and financed by Instituto de Salud Carlos III (ISCIII) – Subdireccion General de Evaluacion and the Fondo Europeo de Desarrollo Regional (FEDER – “Una Manera de Hacer Europa”). We thank Inés Santiuste and the Valdecilla Biobank (PT17/0015/0019), integrated in the Spanish Biobank Network, for their support and collaboration in sample collection and management. The French National Surveillance Network for Creutzfeldt-Jakob disease is supported by Santé Publique France. MDG (UCSF) receives research support from the NIH/NIA (grant R01 AG031189, R56AG055619, R01AG062562) and the Michael J. Homer Family Fund. We thank Megan Casey for assistance with sample collection and management. The National Prion Disease Pathology Surveillance Center in the U.S. is funded by the Centers for Disease Control and Prevention (NU38CK000486). The Austrian Reference Center for Human Prion diseases (ÖRPE) is supported by the Austrian Ministy of Health – Bundesministerium für Soziales, Gesundheit, Pflege und Konsumentenschutz. The Australian National Creutzfeldt-Jakob Disease Registry (ANCJDR) would like to thank all patients and their families for supporting surveillance activities that have allowed participation in the study, as well as their managing physicians: the ANCJDR is supported through funding from the Commonwealth Department of Health and Aged Care.

## Supplementary Material

### Patient recruitment and phenotypes

Samples from patients with prion diseases were provided by specialist or national surveillance centres. A diagnosis of probable or definite sporadic CJD was required according to the contemporary widely accepted epidemiological criteria. These criteria have evolved over time, principally to include the recognition of the importance of MRI brain imaging, and the cerebrospinal fluid Real-Time Quaking Induced Conversion Assay (RT-QuIC) in diagnosis. As there was no restriction on the calendar date of diagnosis many patients were diagnosed using previous versions of diagnostic criteria with similarly high levels of specificity. Ethical approval for research studies was provided by London – Harrow Research Ethics Committee. Clinical duration was defined as the time from first symptom to death (months). Age at clinical onset was defined as the age at the time of the first symptom (years). For more details see Ref 19, main paper.

**Suppl. Fig 1.**
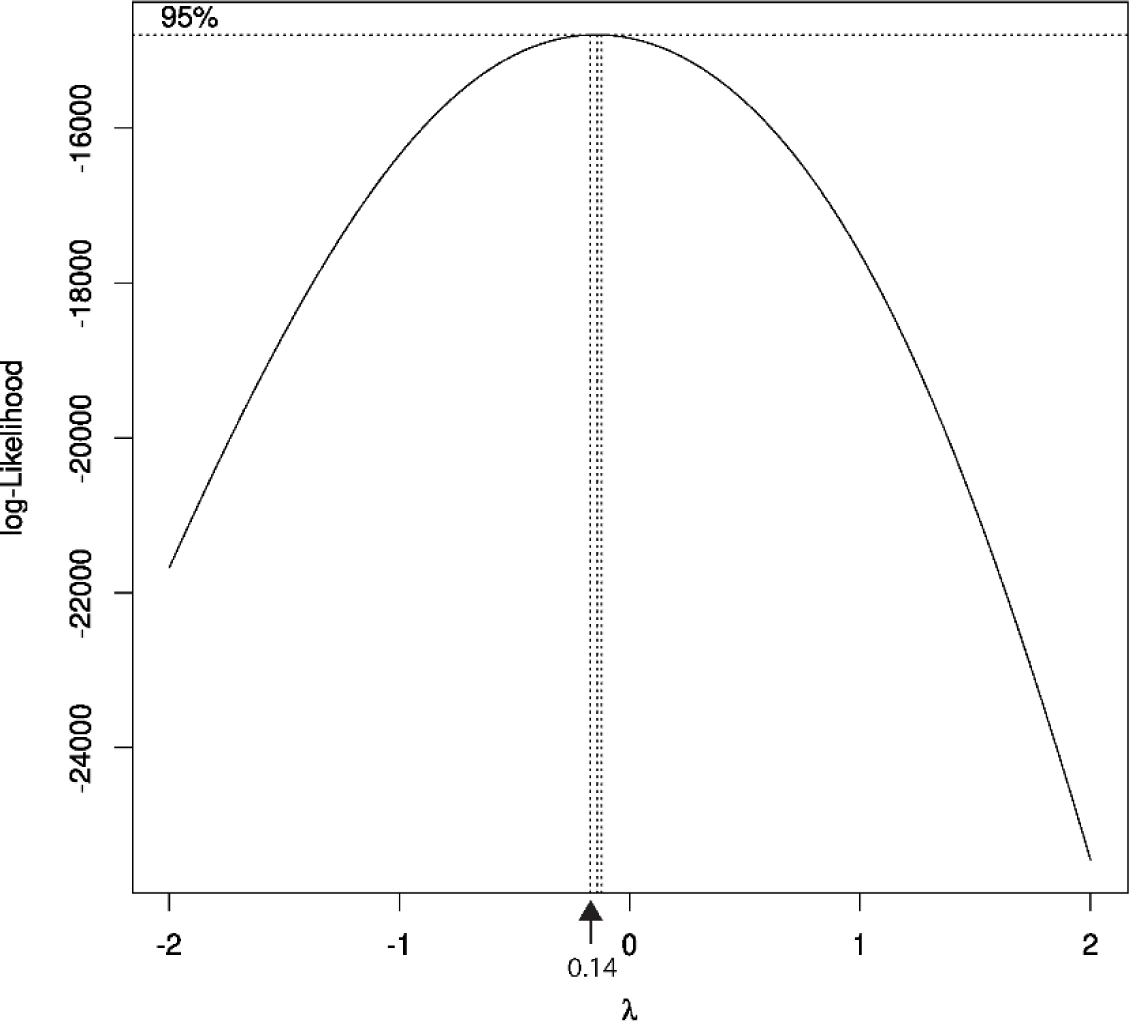
Box-Cox normality plot showing the correlation coefficient (maximum value at -0.14)

**Suppl. Fig 2.**
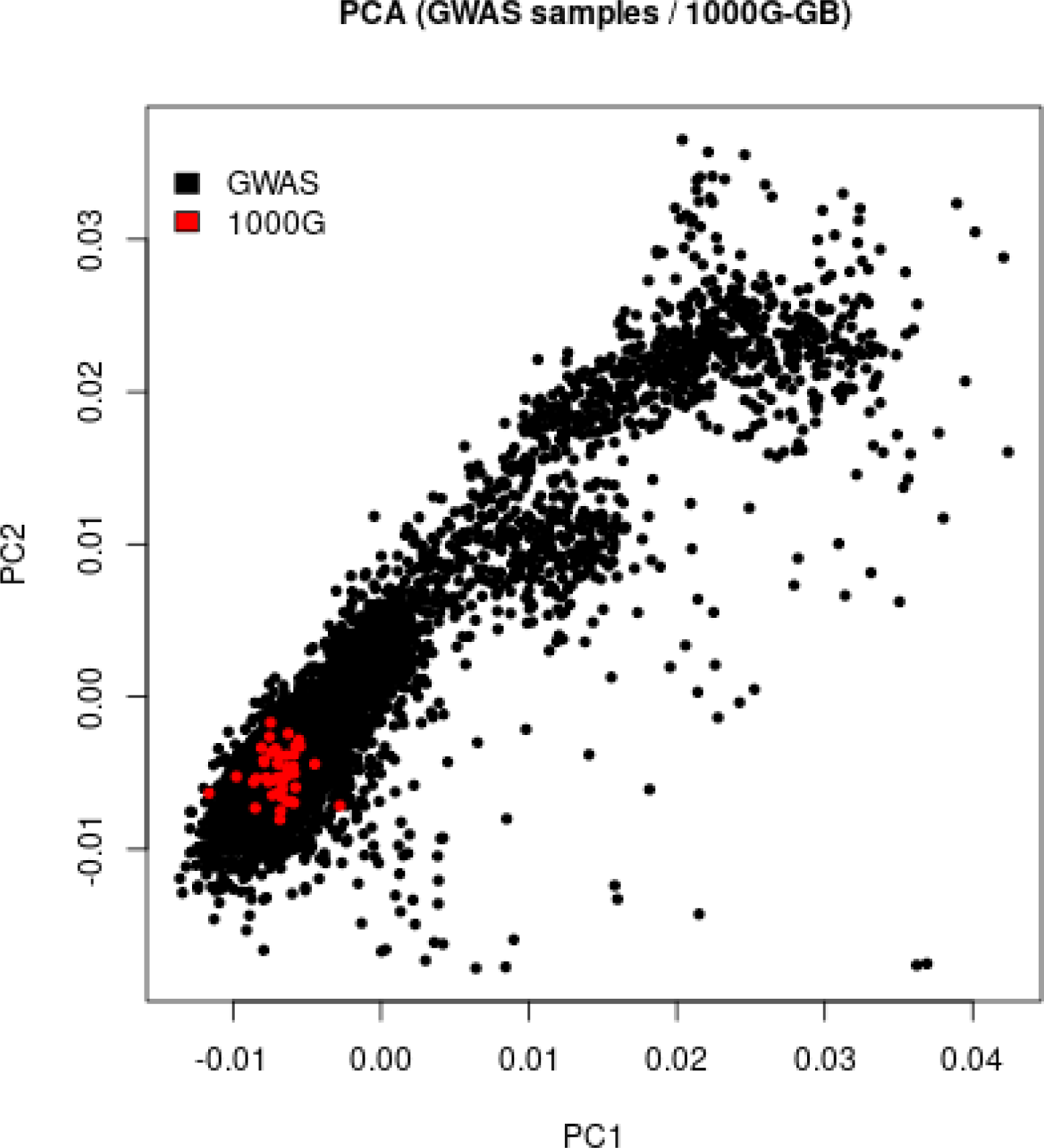
Principal component analysis with first two axes used for exclusion of case and control samples (1000 Genome data used as European ancestry control)

**Suppl. Fig 3.**
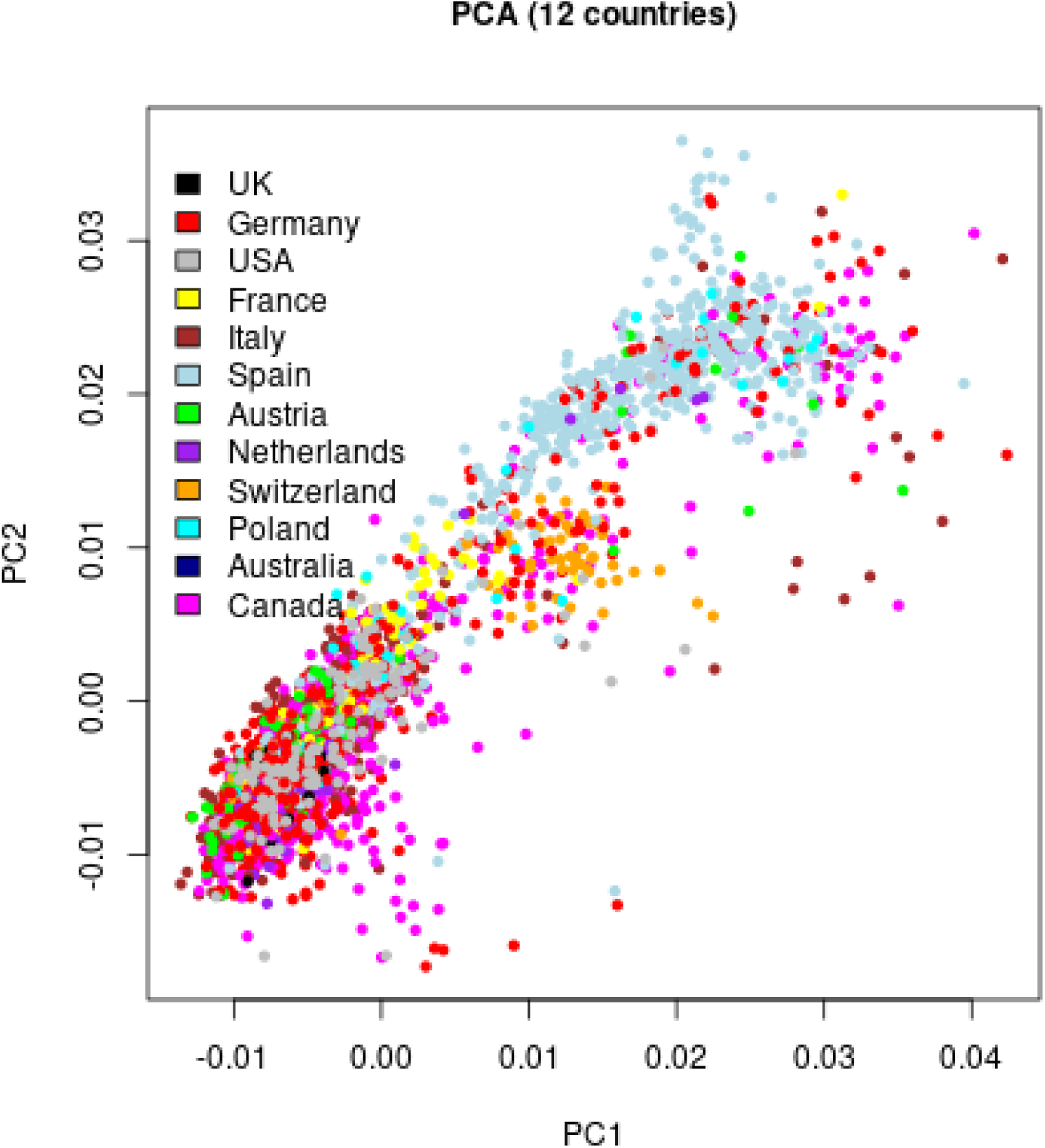
Principal component featuring distribution of samples in terms of country of origin.

**Suppl. Fig 4.**
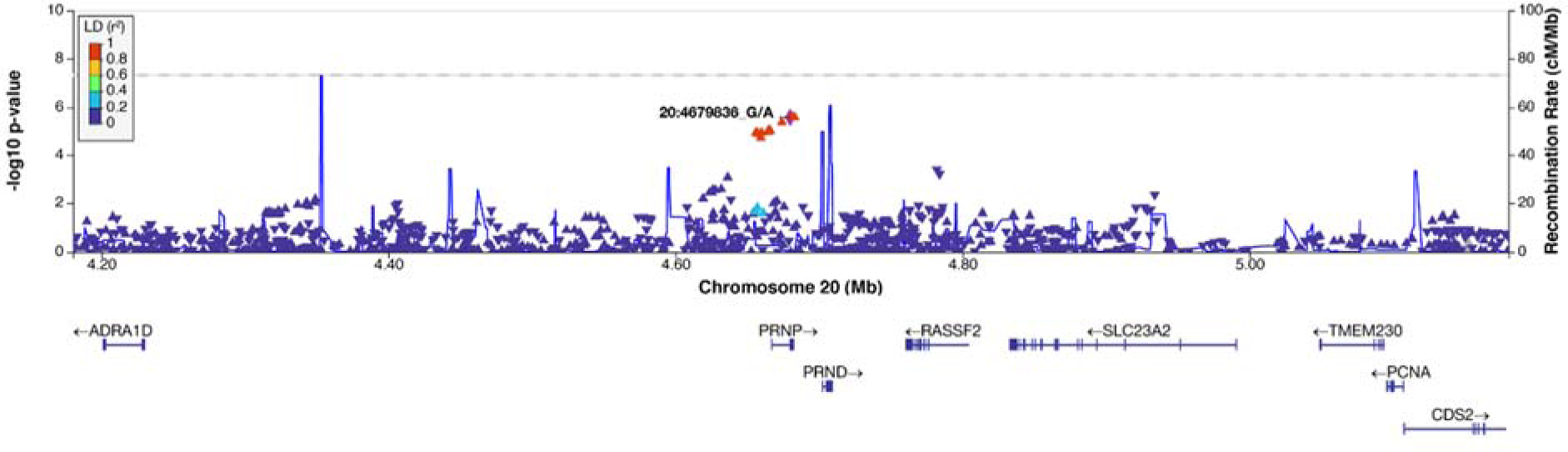
Regional association plot at *PRNP* locus for conditional analysis on SNP rs1799990 with duration as phenotype (heterozygous model)

**Suppl. Fig 5.**
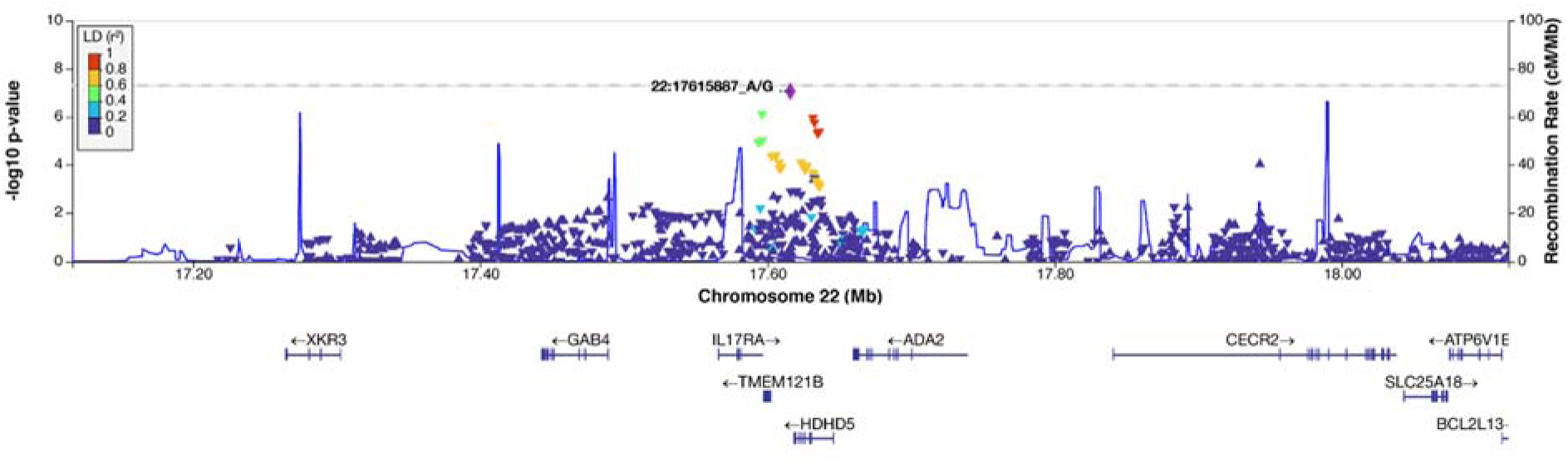
Regional association plot at suggestive locus (*HDHD5*) with duration as phenotype (additive model)

**Suppl. Fig 6.**
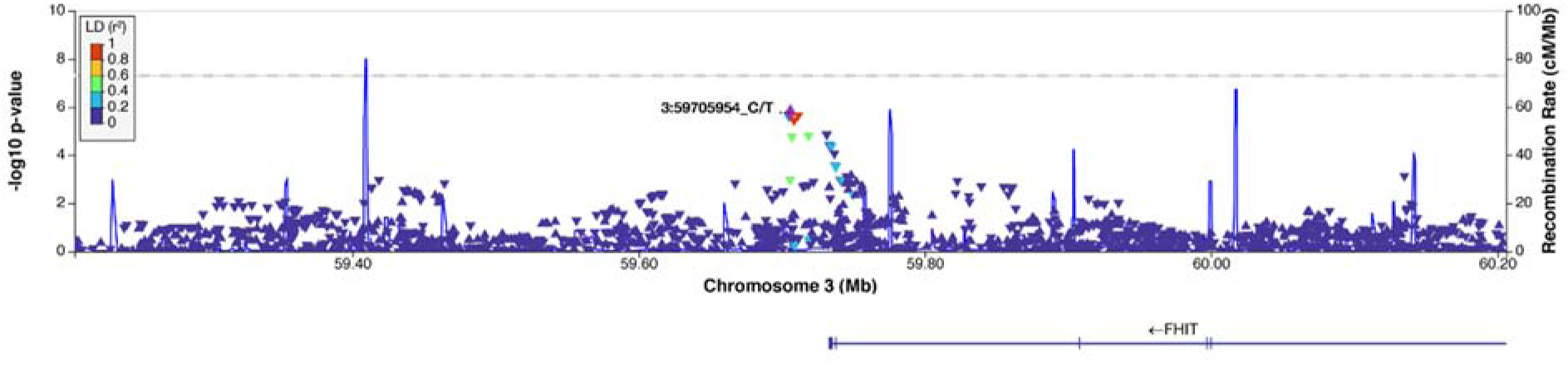
Regional association plot at suggestive *FHIT* locus with duration as phenotype (additive model)

**Suppl. Fig 7.**
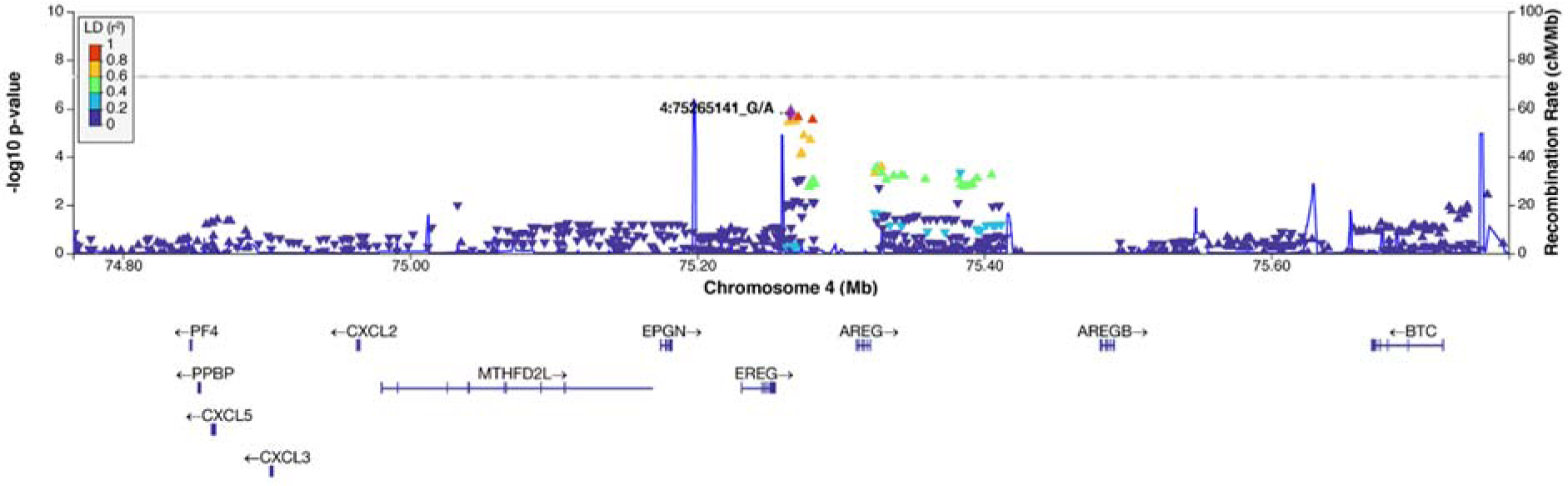
Regional association plot at suggestive *EREG* locus with duration as phenotype (additive model)

**Suppl. Fig 8.**
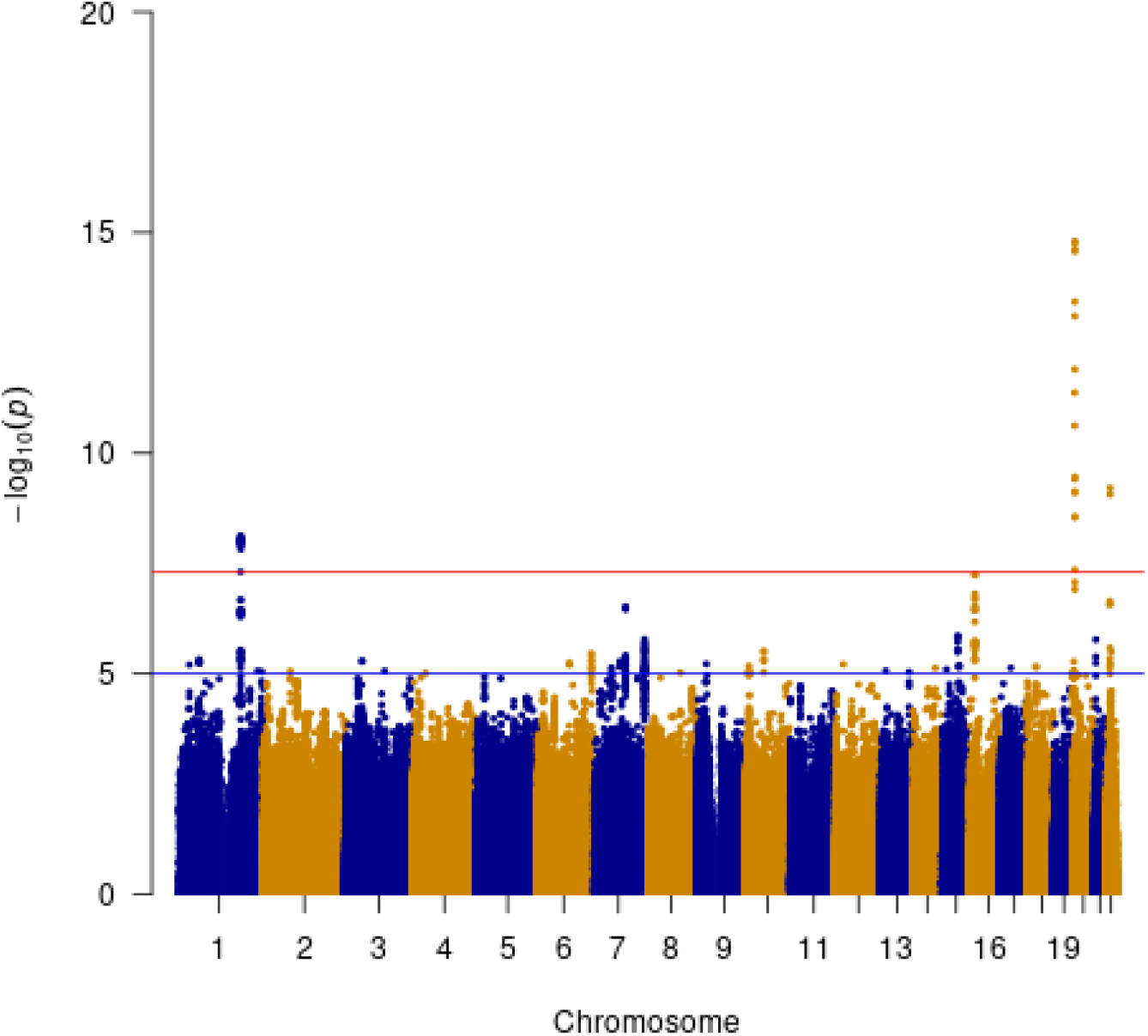
Meta-analysis of case-only and case-control GWAS.

**Suppl. Fig 9.**
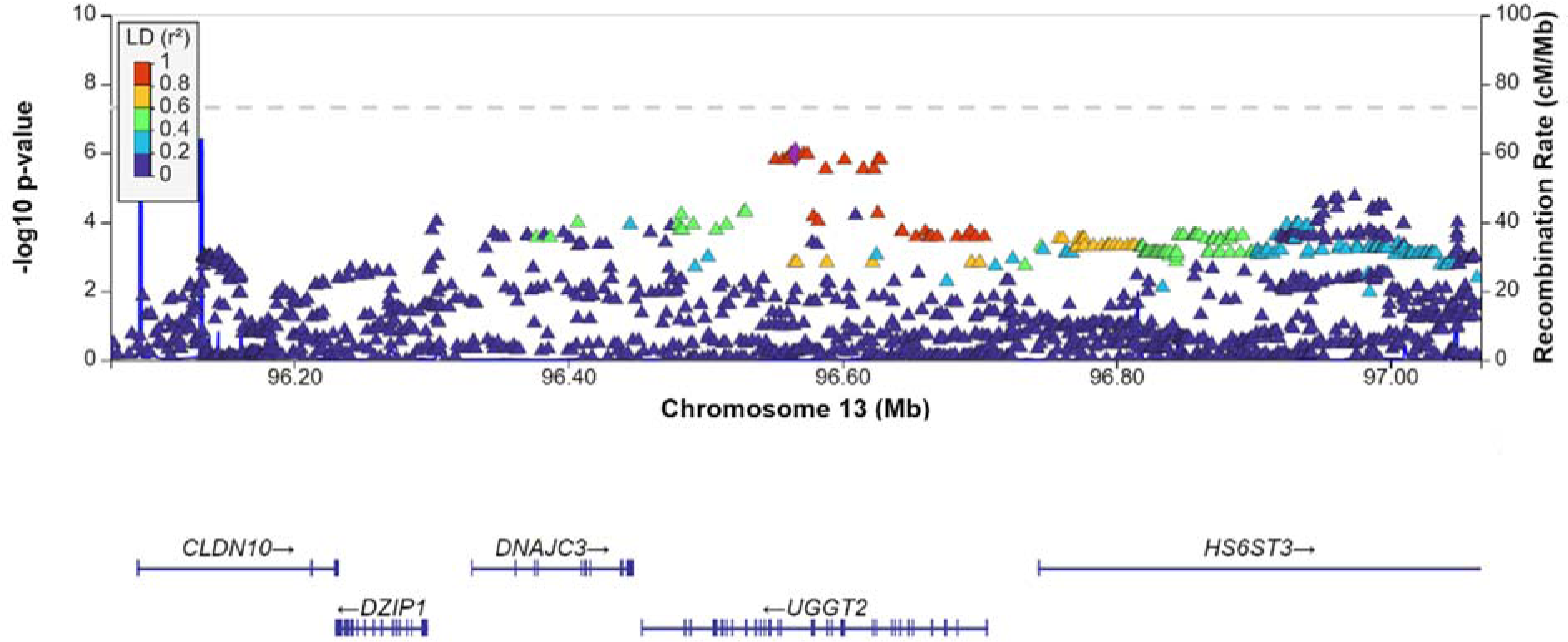
Regional association plot at suggestive *UGGT2* locus with age as phenotype (additive model)

**Suppl. Fig 10.**
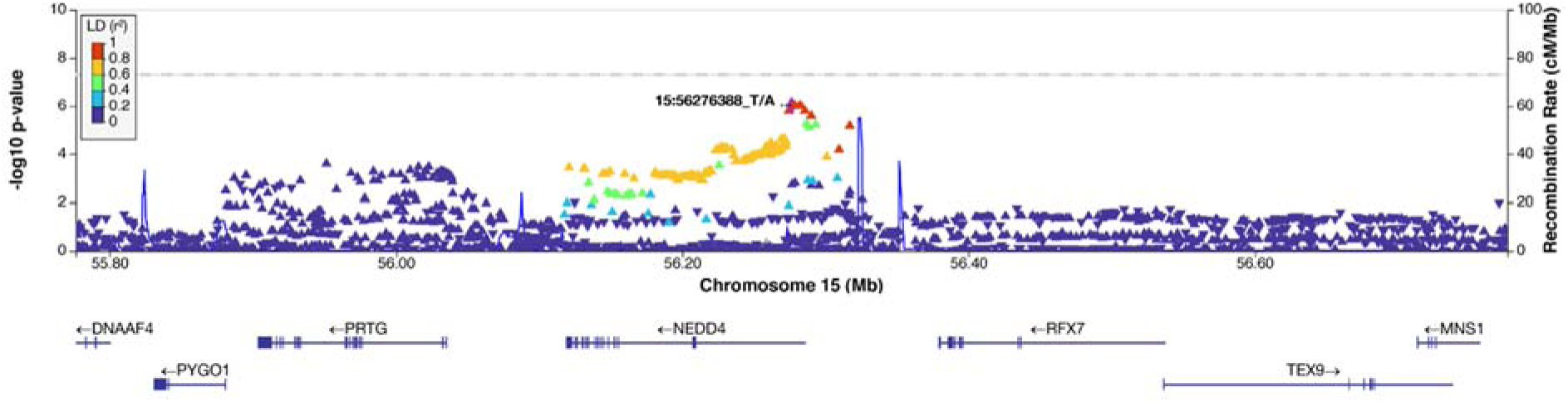
Regional association plot at suggestive *NEDD4* locus with age as phenotype (additive model)

**Suppl. Fig 11.**
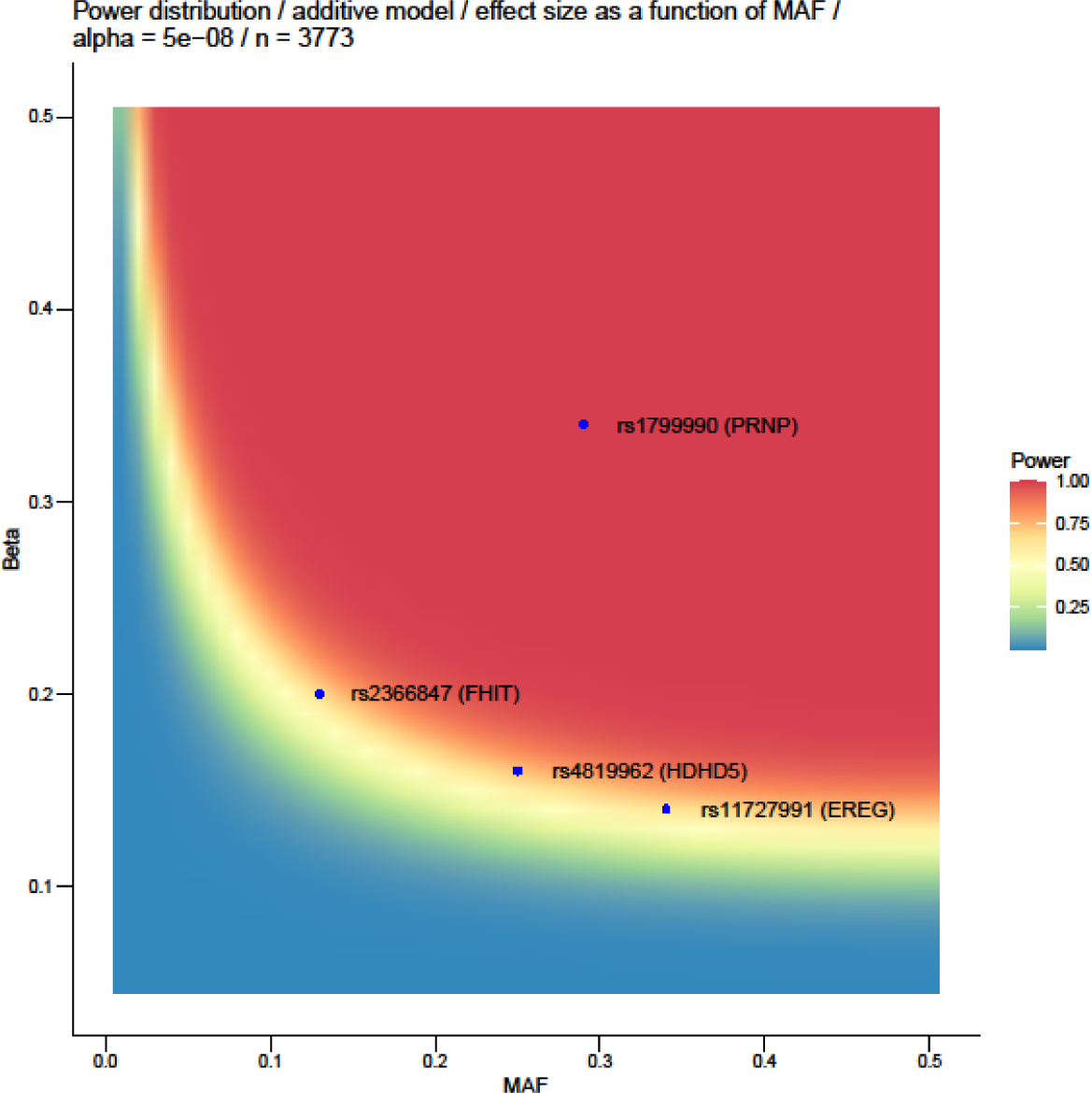
Power analysis indicating the strength of power of *PRNP* lead SNP (rs1799990) and lead SNPs of suggestive hits as described in the Results section based on a range of effect sizes vs. MAF.

**Suppl. Fig 12.**
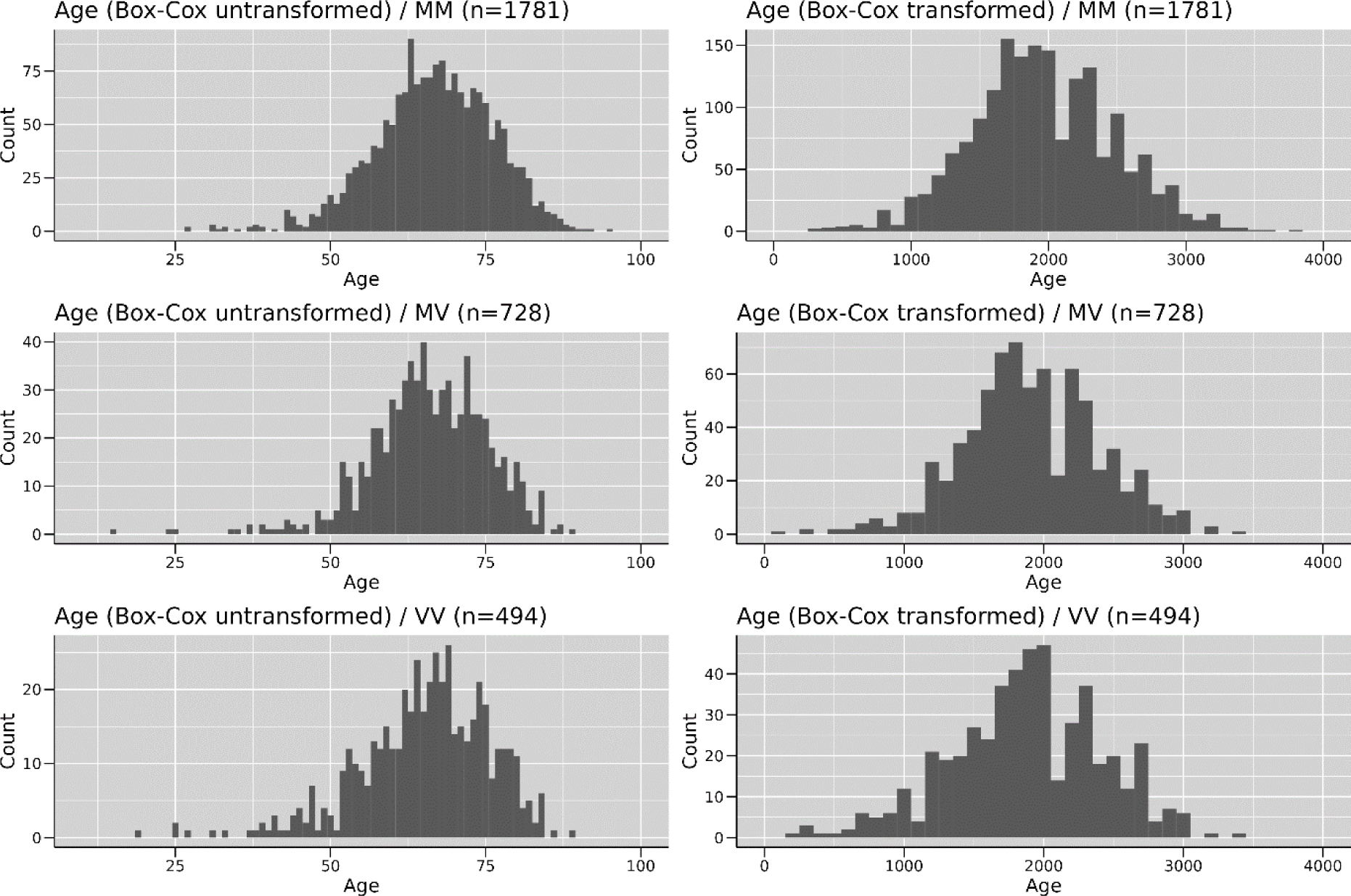
Histograms for phenotype age before and after Box-Cox transformation for codon 129 genotypes MM, MV and VV.

**Suppl. Fig 13.**
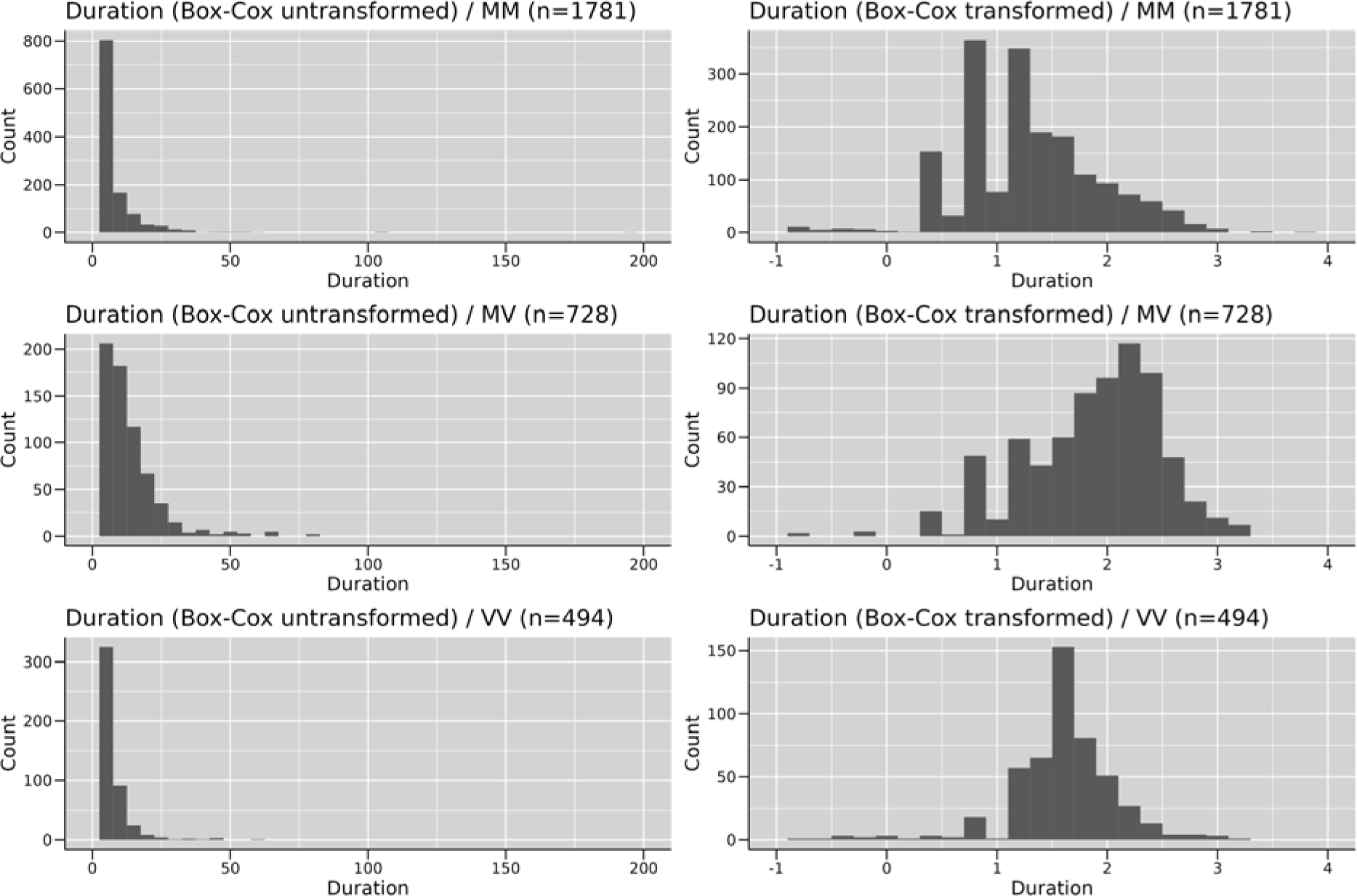
Histograms for phenotype duration before and after Box-Cox transformation for codon 129 genotypes MM, MV and VV.

**Suppl. Table 1a.**
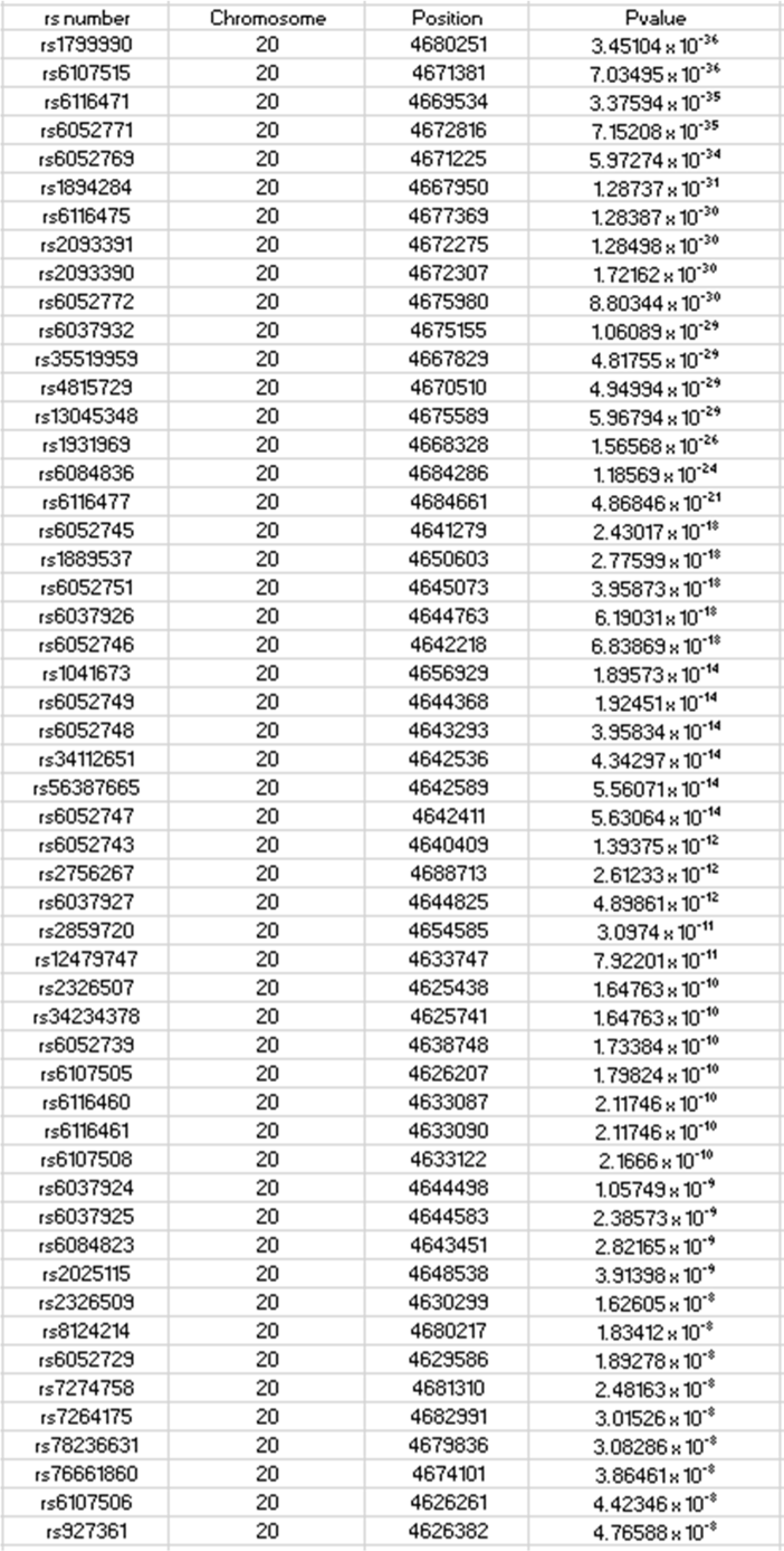
List of 53 significantly associated SNPs (Pvalue < 5×10^-8^)

**Suppl. Table 1b.** List of 51 suggestively associated SNPs (5×10^-8^ > Pvalue < 1×10^-5^)

| rs Number | Chromosome | Position | Pvalue |
| --- | --- | --- | --- |
| rs5994173 | 22 | 17615887 | $8.75 \times 10^{-8}$ |
| rs115391371 | 20 | 4665964 | $1.98 \times 10^{-7}$ |
| rs80244651 | 20 | 4664172 | $2.316 \times 10^{-7}$ |
| rs75916861 | 20 | 4664905 | $2.34 \times 10^{-7}$ |
| rs6037921 | 20 | 4637751 | $4.80 \times 10^{-7}$ |
| rs4819962 | 22 | 17596178 | $7.83 \times 10^{-7}$ |
| rs927359 | 20 | 4626537 | $1.00 \times 10^{-6}$ |
| rs2009160 | 20 | 4626518 | $1.02 \times 10^{-6}$ |
| rs77100741 | 20 | 4657459 | $1.06 \times 10^{-6}$ |
| rs1931970 | 20 | 4622649 | $1.07 \times 10^{-6}$ |
| rs4607002 | 20 | 4625537 | $1.08 \times 10^{-6}$ |
| rs6084817 | 20 | 4625558 | $1.08 \times 10^{-6}$ |
| rs1034862 | 22 | 17631691 | $1.09 \times 10^{-6}$ |
| rs79214721 | 20 | 4655999 | $1.12 \times 10^{-6}$ |
| rs77299769 | 20 | 4656845 | $1.12 \times 10^{-6}$ |
| rs2326506 | 20 | 4625296 | $1.17 \times 10^{-6}$ |
| rs116191575 | 20 | 4659374 | $1.21 \times 10^{-6}$ |
| rs115148957 | 20 | 4659375 | $1.21 \times 10^{-6}$ |
| rs6052715 | 20 | 4624725 | $1.25 \times 10^{-6}$ |
| rs6037913 | 20 | 4624176 | $1.27 \times 10^{-6}$ |
| rs6052717 | 20 | 4625001 | $1.27 \times 10^{-6}$ |
| rs2326504 | 20 | 4625108 | $1.27 \times 10^{-6}$ |
| rs2326505 | 20 | 4625139 | $1.27 \times 10^{-6}$ |
| rs927358 | 20 | 4626704 | $1.33 \times 10^{-6}$ |
| rs114970370 | 20 | 4659887 | $1.40 \times 10^{-6}$ |
| rs11727991 | 4 | 75265141 | $1.55 \times 10^{-6}$ |
| rs12624635 | 20 | 4636274 | $1.68 \times 10^{-6}$ |
| rs4819560 | 22 | 17632680 | $1.72 \times 10^{-6}$ |
| rs2366847 | 3 | 59705954 | $1.76 \times 10^{-6}$ |
| rs61825724 | 1 | 204035418 | $2.10 \times 10^{-6}$ |
| rs1823230 | 3 | 59711526 | $2.20 \times 10^{-6}$ |
| rs28465052 | 4 | 75270190 | $2.22 \times 10^{-6}$ |
| rs11087649 | 20 | 4636693 | $2.33 \times 10^{-6}$ |
| rs73092458 | 3 | 59704710 | $2.47 \times 10^{-6}$ |
| rs80243481 | 15 | 54244286 | $2.61 \times 10^{-6}$ |
| rs6116466 | 20 | 4637168 | $2.63 \times 10^{-6}$ |
| rs28551637 | 4 | 75280372 | $2.82 \times 10^{-6}$ |
| rs28873375 | 4 | 75267360 | $3.30 \times 10^{-6}$ |
| rs1494882 | 4 | 75268407 | $3.37 \times 10^{-6}$ |
| rs2047193 | 4 | 75268730 | $3.37 \times 10^{-6}$ |
| rs9994668 | 4 | 75263409 | $3.56 \times 10^{-6}$ |
| rs1447803 | 3 | 59708541 | $3.58 \times 10^{-6}$ |
| rs4819972 | 22 | 17635858 | $4.21 \times 10^{-6}$ |
| rs1005326 | 22 | 17634971 | $4.87 \times 10^{-6}$ |
| rs12061308 | 1 | 37717641 | $6.29 \times 10^{-6}$ |
| rs13282211 | 8 | 8760138 | $6.33 \times 10^{-6}$ |
| rs6107507 | 20 | 4633115 | $6.41 \times 10^{-6}$ |
| rs117151717 | 22 | 44213609 | $8.21 \times 10^{-6}$ |
| rs151328756 | 12 | 26430376 | $8.62 \times 10^{-6}$ |
| rs11264006 | 1 | 37725897 | $9.59 \times 10^{-6}$ |
| rs1654 | 22 | 17596388 | $9.77 \times 10^{-6}$ |

**Suppl. Table 2.** Genetic correlation using LDSC with a genetic correlation of 0.1467.

|  |  |
| --- | --- |
| Trait 1 | Case-control |
| Trait 2 | Case-only |
| Genetic correlation (rg) | 0.1467 |
| Standard error of rg | 0.5457 |
| 95% CI interval | 1.21/-0.92 |
| Zscore for rg | 0.2688 |
| Pvalue for rg | 0.7881 |
| Heritability for trait2 (h1) | 0.1819 |
| Heritability for trait2 (h2) | 0.0895 |

**Suppl. Table 3.** Top 10 genes identified by MAGMA involved in sphingolipid and intracellular trafficking pathways (including genome-wide significant SNPs) with age as phenotype. Green rectangles indicate genes involved in neurological diseases (PrD = Prion Disease, PD = Parkinson’s Disease, AD= Alzheimer’s Disease, ALS = Amyotrophic Lateral Sclerosis; HD = Huntington’s Disease, FTD = Frontotemporal Dementia) (NSNPS = number of SNPs annotated to a gene; N = number of samples; ZSTAT = Z-score for the gene, based on its p-value)

| GENE | NCBI GENE ID | CHR | START | STOP | NSNPS | N | ZSTAT | Pvalue | Pvalue (Bonferroni corr.) | PrD | PD | AD | ALS | HD | FTD | Class |
| --- | --- | --- | --- | --- | --- | --- | --- | --- | --- | --- | --- | --- | --- | --- | --- | --- |
| UGGT2 | 55757 | 13 | 96453834 | 96705736 | 389 | 3773 | 4.02 | $2.87 \times 10^{-5}$ | $2.64 \times 10^{-3}$ | | | | | | | Sphingolipid Metabolism |
| CD2AP | 23607 | 6 | 47445482 | 47594999 | 327 | 3773 | 1.74 | $4.14 \times 10^{-2}$ | 1 | | | | | | | Intracellular trafficking |
| SORT1 | 6272 | 1 | 109852187 | 109940563 | 96 | 3773 | 1.72 | $4.30 \times 10^{-2}$ | 1 | | | | | | | Intracellular trafficking |
| CERS5 | 91012 | 12 | 50523581 | 50561154 | 48 | 3773 | 1.43 | $7.60 \times 10^{-2}$ | 1 | | | | | | | Sphingolipid Metabolism |
| VPS29 | 51699 | 12 | 110929330 | 110939916 | 17 | 3773 | 1.31 | $9.58 \times 10^{-2}$ | 1 | | | | | | | Intracellular trafficking |
| CTSB | 1508 | 8 | 11700033 | 11725646 | 107 | 3773 | 1.22 | $1.12 \times 10^{-1}$ | 1 | | | | | | | Intracellular trafficking |
| EPHA4 | 2043 | 2 | 222282747 | 222437010 | 383 | 3773 | 1.17 | $1.22 \times 10^{-1}$ | 1 | | | | | | | Intracellular trafficking |
| CERS2 | 29956 | 1 | 150937649 | 150947479 | 21 | 3773 | 1.04 | $1.50 \times 10^{-1}$ | 1 | | | | | | | Sphingolipid Metabolism |
| C9orf72 | 203228 | 9 | 27546543 | 27573864 | 91 | 3773 | 1.00 | $1.58 \times 10^{-1}$ | 1 | | | | | | | Intracellular trafficking |
| RAB10 | 10890 | 2 | 26256729 | 26360323 | 145 | 3773 | 0.95 | $1.72 \times 10^{-1}$ | 1 | | | | | | | Intracellular trafficking |

**Suppl. Table 4.** Top 10 genes identified by MAGMA involved in sphingolipid and intracellular trafficking pathways (including genome-wide significant SNPs) with duration as phenotype. Green rectangles indicate genes involved in neurological diseases (PrD = Prion Disease, PD = Parkinson’s Disease, AD= Alzheimer’s Disease, ALS = Amyotrophic Lateral Sclerosis; HD = Huntington’s Disease, FTD = Frontotemporal Dementia) (NSNPS = number of SNPs annotated to a gene; N = number of samples; ZSTAT = Z-score for the gene, based on its p-value)

| GENE | NCBI GENE ID | CHR | START (hg19) | STOP (hg19) | NSNPS | N | ZSTAT | Pvalue | Pvalue (Bonferroni corr.) | PrD | PD | AD | ALS | HD | FTD | Class |
| --- | --- | --- | --- | --- | --- | --- | --- | --- | --- | --- | --- | --- | --- | --- | --- | --- |
| UGGT2 | 55757 | 13 | 96453834 | 96705736 | 389 | 3773 | 2.39 | $8.50 \times 10^{-3}$ | 0.78 | | | | | | | Sphingolipid Metabolism |
| SGPL1 | 8879 | 10 | 72575704 | 72640946 | 180 | 3773 | 2.13 | $1.66 \times 10^{-2}$ | 1 | | | | | | | Sphingolipid Metabolism |
| UGT8 | 7368 | 4 | 115519611 | 115598202 | 111 | 3773 | 2.06 | $1.96 \times 10^{-2}$ | 1 | | | | | | | Sphingolipid Metabolism |
| SOAT1 | 6646 | 1 | 179262849 | 179327815 | 239 | 3773 | 1.85 | $3.19 \times 10^{-2}$ | 1 | | | | | | | Cholesterol Homeostasis |
| C9orf72 | 203228 | 9 | 27546543 | 27573864 | 91 | 3773 | 1.81 | $3.50 \times 10^{-2}$ | 1 | | | | | | | Intracellular trafficking |
| NAGLU | 4669 | 17 | 40687951 | 40696467 | 5 | 3773 | 1.76 | $3.90 \times 10^{-2}$ | 1 | | | | | | | GAG Metabolism |
| CERS2 | 29956 | 1 | 150937649 | 150947479 | 21 | 3773 | 1.42 | $7.81 \times 10^{-2}$ | 1 | | | | | | | Sphingolipid Metabolism |
| UGCG | 7357 | 9 | 114659206 | 114697649 | 141 | 3773 | 1.19 | $1.16 \times 10^{-1}$ | 1 | | | | | | | Sphingolipid Metabolism |
| PTDSS1 | 9791 | 8 | 97274167 | 97346774 | 90 | 3773 | 1.17 | $1.22 \times 10^{-1}$ | 1 | | | | | | | Sphingolipid Metabolism |
| VPS26A | 9559 | 10 | 70883908 | 70932617 | 98 | 3773 | 1.14 | $1.28 \times 10^{-1}$ | 1 | | | | | | | Intracellular trafficking |

**Suppl. Table 5a.**
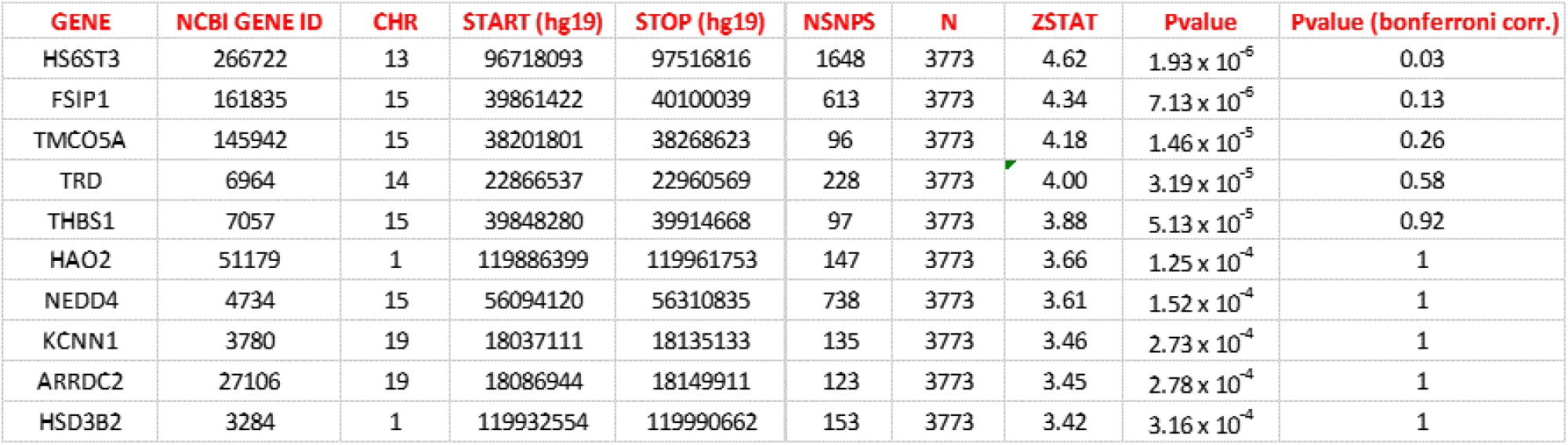
Top 10 genes identified by MAGMA (standalone) gene analysis (including genome-wide significant SNPs) with age as phenotype. (NSNPS = number of SNPs annotated to a gene; N = number of samples; ZSTAT = Z-score for the gene, based on its p-value)

**Suppl. Table 5b.**
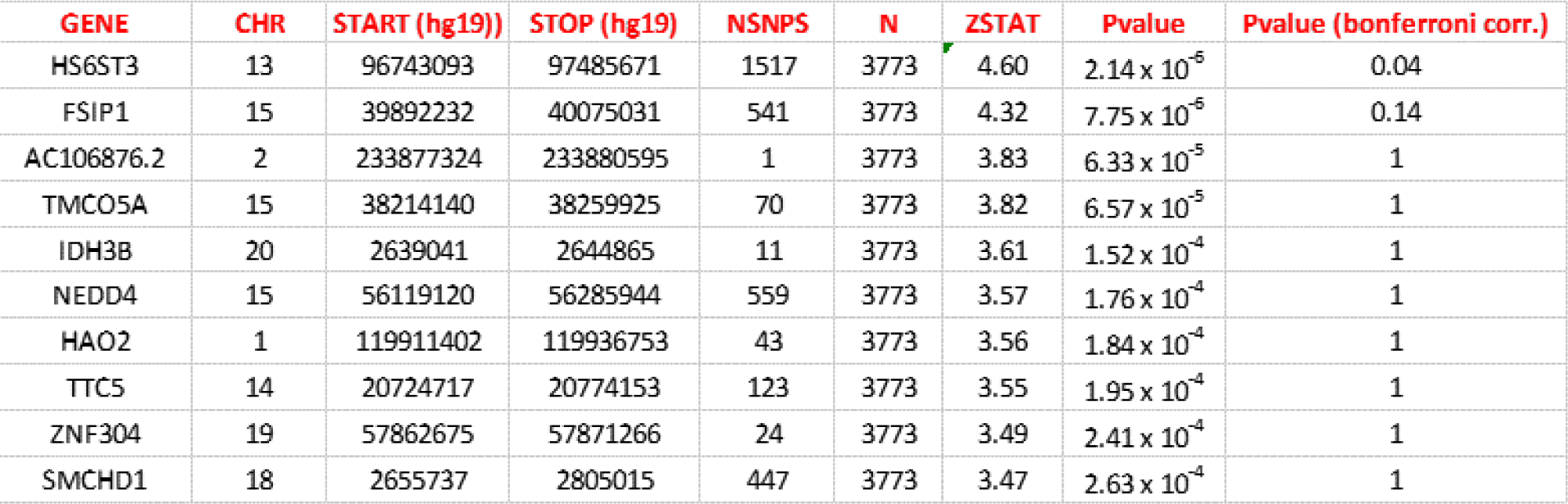
Top 10 genes identified by FUMA gene analysis (including genome-wide significant SNPs) with age as phenotype. (NSNPS = number of SNPs annotated to a gene; N = number of samples; ZSTAT = Z-score for the gene, based on its p-value)

**Suppl. Table 5c.**
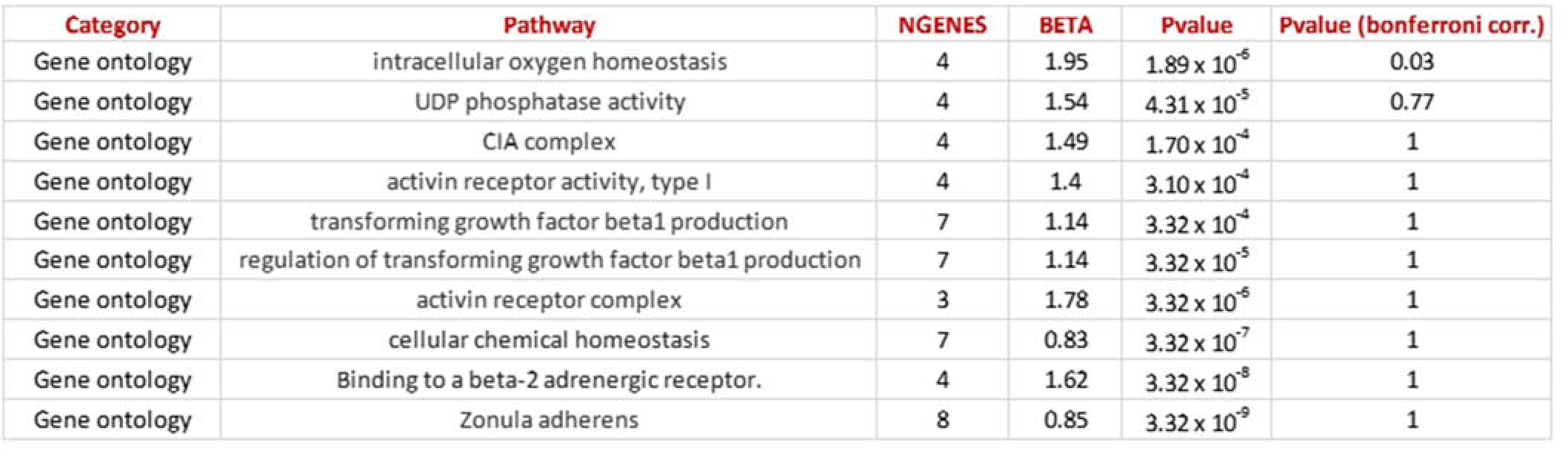
Top 10 pathways identified by MAGMA (standalone) gene-set analysis (including genome-wide SNPs) using age as phenotype. (NGENES = number of genes in the gene-set dataset; BETA = regression coefficient of the gene set)

**Suppl. Table 5d.** Top 10 pathways identified by FUMA gene-set analysis (including genome-wide SNPs) using age as phenotype. (NGENES = number of genes in the gene-set dataset; BETA = regression coefficient of the gene set)

| Category | Pathway | NGENES | BETA | Pvalue | Pvalue (bonferroni corr.) |
| --- | --- | --- | --- | --- | --- |
| Curated gene sets | reactome_activation_of_c3_and_c5 | 6 | 1.40 | $1.87 \times 10^{-5}$ | 0.29 |
| Curated gene sets | pasqualucci_lymphoma_by_gc_stage_up | 263 | 0.20 | $3.94 \times 10^{-5}$ | 0.61 |
| Gene ontology | go_rna_adenylyltransferase_activity | 4 | 1.28 | $1.01 \times 10^{-4}$ | 1 |
| Gene ontology | go_nuclear_outer_membrane_endoplasmic_reticulum_membrane_network | 995 | 0.10 | $1.12 \times 10^{-4}$ | 1 |
| Gene ontology | go_endoplasmic_reticulum_part | 1231 | 0.09 | $1.97 \times 10^{-4}$ | 1 |
| Gene ontology | go_regulation_of_phosphate_transport | 8 | 1.05 | $2.95 \times 10^{-4}$ | 1 |
| Gene ontology | go_renal_absorption | 14 | 0.75 | $3.08 \times 10^{-4}$ | 1 |
| Curated gene sets | reactome_complement_cascade | 48 | 0.46 | $3.08 \times 10^{-4}$ | 1 |
| Curated gene sets | reactome_initial_triggering_of_complement | 18 | 0.79 | $3.68 \times 10^{-4}$ | 1 |
| Curated gene sets | biocarta_comp_pathway | 17 | 0.79 | $3.94 \times 10^{-4}$ | 1 |

